# Small-molecule metabolome identifies potential therapeutic targets against COVID-19

**DOI:** 10.1101/2021.06.18.21259150

**Authors:** Sean M. P. Bennet, Martin Kaufmann, Kaede Takami, Calvin Sjaarda, Katya Douchant, Emily Moslinger, Henry Wong, David E. Reed, Anne K. Ellis, Stephen Vanner, Robert I. Colautti, Prameet M. Sheth

## Abstract

**Background:** Respiratory viruses are transmitted and acquired via the nasal mucosa, and thereby may influence the nasal metabolome composed of biochemical products produced by both host cells and microbes. Studies of the nasal metabolome demonstrate virus-specific changes that sometimes correlate with viral load and disease severity. Here, we evaluate the nasopharyngeal metabolome of COVID-19 infected individuals and report several small molecules that may be used as potential therapeutic targets. Specimens were tested by qRT-PCR with target primers for three viruses: Influenza A (INFA), respiratory syncytial virus (RSV), and SARS-CoV-2, along with asymptomatic controls. The nasopharyngeal metabolome was characterized using an LC-MS/MS-based small-molecule screening kit capable of quantifying 141 analytes. A machine learning model identified 28 discriminating analytes and correctly categorized patients with a viral infection with an accuracy of 96% (R^2^=0.771, Q^2^=0.72). A second model identified 5 analytes to differentiate COVID19-infected patients from those with INFA or RSV with an accuracy of 85% (R^2^=0.442, Q^2^=0.301). Specifically, LysoPCaC18:2 concentration was significantly increased in COVID19 patients (P< 0.0001), whereas beta-hydroxybutyric acid, Met SO, succinic acid, and carnosine concentrations were significantly decreased (P< 0.0001). This study demonstrates that COVID19 infection results in a unique NP metabolomic signature with carnosine and LysoPCaC18:2 as potential therapeutic targets.

**Significance Statement:** Efforts to elucidate how SARS-CoV-2 interacts with the host has become a global priority. To identify biomarkers for potential therapeutic interventions, we used a targeted metabolomics approach evaluating metabolite profiles in the nasal mucosa of COVID-19 patients and compared metabolite profiles to those of other respiratory viruses (influenza A, RSV). We identified a COVID-19-specific signature characterized by changes to LysoPCaC18:2, beta-hydroxybutyric acid, Met SO, succinic acid, and carnosine. Carnosine is a promising potential target against SARS-CoV-2 as it has been shown to interfere with binding of SARS-CoV-2 to the ACE2 receptor. This study provides compelling evidence for the use of metabolomics as an avenue for the identification of novel drug targets for viral respiratory infections in the nasopharynx.

## Introduction

COVID-19 represents one of the greatest public health challenges of the 21st century. Unlike most respiratory viruses, SARS-CoV-2 has a longer incubation period and infected individuals present with a spectrum of symptoms ranging from asymptomatic to severe clinical disease requiring hospitalization. The majority of SARS-CoV-2 infections occur via the nasal mucosa (1). Understanding the host-pathogen interactions in the nasal mucosa may provide valuable insight into the identification of novel therapeutic targets. These targets may be used to interrupt the acquisition and limit disease progression of SARS-CoV-2. We thus examined if the nasal metabolomic profile for COVID-19 was distinct from those of other respiratory viruses, and whether examining the metabolome of the nasopharynx (NP) would provide insight into host-pathogen interactions in the nasal mucosa.

Previous studies involving the nasal metabolome in individuals infected with respiratory viruses, including rhinovirus (RV) and respiratory syncytial virus (RSV), reported that the nasal metabolome was virus-specific, despite indistinguishable clinical presentations in infected individuals (2). Furthermore, the concentrations of specific nasal metabolites positively correlated with viral load and disease severity and predicted the need for positive pressure ventilation in patients with a high degree of sensitivity and specificity (84% and 86%, respectively)(3). The predominant changes in the nasal metabolome observed in response to respiratory viruses were identified to be host-derived, although some metabolite concentrations correlated with colonization with *Haemophilus influenzae, Streptococcus pneumoniae* and *Moraxella catarrhalis* (2). These studies suggest that evaluating metabolic signatures in the nasopharynx of COVID-19 patients compared to other respiratory viruses may provide insight into important host-mediated antiviral responses, further elucidate changes that may be occurring in the nasal microbial environment and potentially identify new therapeutic targets against COVID-19.

In this study we hypothesized that SARS-CoV-2 induces a characteristic change to the nasal metabolome of patients and that could be used to both identify metabolites important in pathogenicity and potential therapeutic targets. We thus implemented a targeted LC-MS/MS metabolomics approach to (i) characterize small-molecule profiles in viral transport media from NP swabs of patients infected with INFA, RSV or COVID-19 and asymptomatic controls; (ii) identify COVID-19 specific metabolite patterns; and (iii) explore potential therapeutic pathways based on significant metabolites identified by a supervised machine learning model.

## Results

A total of 210 individuals were included in this study, comprising four patient groups: 44 asymptomatic controls (AC), 55 patients positive for SARS-CoV-2 (COV), 55 patients positive for INFA and 56 positive for RSV (Table 1). PCR Cycle Threshold (CT) values were used as a surrogate for viral load (VL) and samples were stratified according to CT’s of 30-35 (low VL), 25-30 (intermediate VL) and < 24.9 (high VL).

**Table 1:** Patient demographics.

|  | Patient Group |  |  |  |
| --- | --- | --- | --- | --- |
|  | SARS-COV-2 (COV) | Influenza A (INFA) | Respiratory Syncytial Virus (RSV) | Asymptomatic Controls |
| **N** | 55 | 55 | 56 | 44 |
| **Year of Collection (range in Months)** | 2020<br>(Jan - Apr) | 2019 - 2020<br>(Dec-Mar) | 2019 - 2020<br>(Dec-Mar) | 2020<br>(Sept) |
| **Median Age (Range)** | 55 years<br>(20-85 years) | 62 years<br>(27 days - 94 years) | 16 months<br>(21 days - 91 years) | 24 years<br>(19 - 43 years) |
| **Sex (%)** | M 27%<br>F 42%<br>n/a 31% | M 54%<br>F 44%<br>n/a 2% | M 40%<br>F 46%<br>n/a 14% | M 25%<br>F 75% |
| **Median CT<sup>1</sup> (Range)** | 26.2<br>(17.76 - 37.24) | 28.6<br>(21.43 - 38.6) | 26.3<br>(18.54 - 36.87) | N/A |

<sup>1</sup> PCR cycle threshold values (CT)

We studied the nasopharyngeal metabolome in patients who underwent standard-of-care or screen testing for respiratory infection. Viral transport media (VTM) from clinical samples were analyzed using a targeted, small-molecule screening kit (TMIC Prime) capable of quantifying 141-analytes over six chemical classes using a combination of LC-MS/MS and flow-injection analysis-MS/MS (Supplementary Fig S1). Upon examination of individual analytes, 44 chemical species exhibited at least double the concentration in patient samples as compared with blank VTM, comprising amino acids (N=17), organic acids (N=4), biogenic amines (N=14) acylcarnitines (N=1) and lipids (N=8). Similarly, the feature selection step of our machine learning pipeline identified a subset of 28 metabolites that differed significantly between infected patients and asymptomatic controls, and 5 metabolites that distinguished SARS-CoV-2 from the other respiratory diseases. Combining these yielded 30 unique metabolites that we prioritized for multivariate analysis, including amino acids (N=15), organic acids (N=4), acylcarnitines (N=1), lipids (N=4), biogenic amines (N=5) and total hexoses (N=1) (Supplementary Table S1). Multivariate modelling of metabolite profiles scaled to blank VTM using partial-least discriminant analysis (PLS-DA) revealed separation of the four patient groups (Figure 1A, B). A second model using orthogonal partial least squares discriminant analysis (OPLS-DA) focused on differences between asymptomatic patients and those with respiratory illness (INFA, RSV or COVID19) (Fig. 1C). Using half of the data for training and the other half for testing, this model had an accuracy of 96%, a sensitivity of 98% and specificity of 86% (R^2^=0.771, Q^2^=0.72) in differentiating between groups (Fig. 1E). A third model compared COVID19 patients and those with other respiratory illnesses (INFA or RSV) (Fig. 1D) Using the same cross-validation analysis, this model distinguished patients with an accuracy of 85%, sensitivity of 74% and specificity of 90 % (R^2^=0.442, Q^2^=0.301) (Fig. 1F).

**Figure 1:**
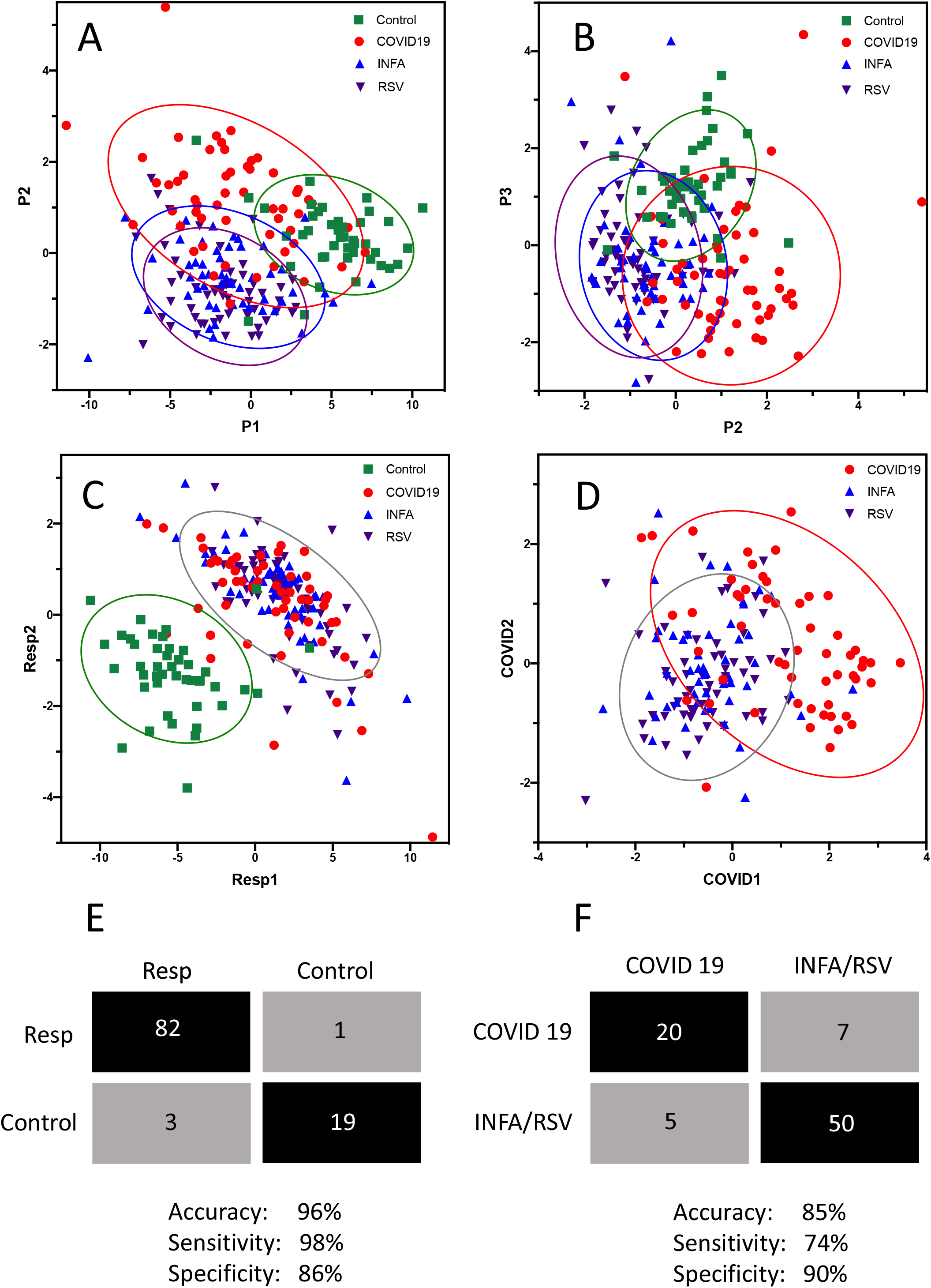
Multivariate analysis and classification of patients with respiratory illness based on metabolite profiles. Supervised partial least squares discriminant analysis was used to plot analyte profiles in VTM from clinical nasopharyngeal swabs scaled to control VTM. Plots of components 1 and 2 (A) and 1 and 3 (B) are shown where optimal separation of patient groups was observed. Orthogonal partial least squares discriminant analysis was used to plot analyte profiles among patient groups. In panel C, all patients with a respiratory illness were grouped into a single category and compared to asymptomatic subjects. In panel D, COVID19 patients were compared to all other patients with influenza A and RSV were tr into a single category. The 95% confidence region is circled for each category. Confusion matrices based on test/train cohorts using 50% of the data are shown in panels E and F from which the accuracy, sensitivity and specificity of identifying patients with a respiratory infection in general (E) or patients with COVID19 among patients with respiratory illness (F) was determined.

OPLS-DA loadings for the 30 unique metabolites that differed significantly among patient groups and controls are shown in Figure 2A. Twenty-eight analytes exhibited increased concentrations in patients with respiratory infections as compared with asymptomatic controls, including amino acids, lipids, organic acids, and biogenic amines. Most importantly, a smaller subset of analytes was observed to be specifically increased (LysoPCaC18:2) or decreased (MetSO, beta hydroxy-butyric acid, carnosine, and succinic acid) in COVID19 patients as compared with INFA or RSV patients (Figure 2B). Interestingly, carnosine and succinic acid were not found to be important factors in differentiating all respiratory patients from controls (Figure 2A and 2B). Despite their ability to distinguish among patient groups, none of these five metabolites correlated significantly with VL (qRT-PCR CT) for any of the three respiratory viruses (Supplementary Fig. S2).

**Figure 2:**
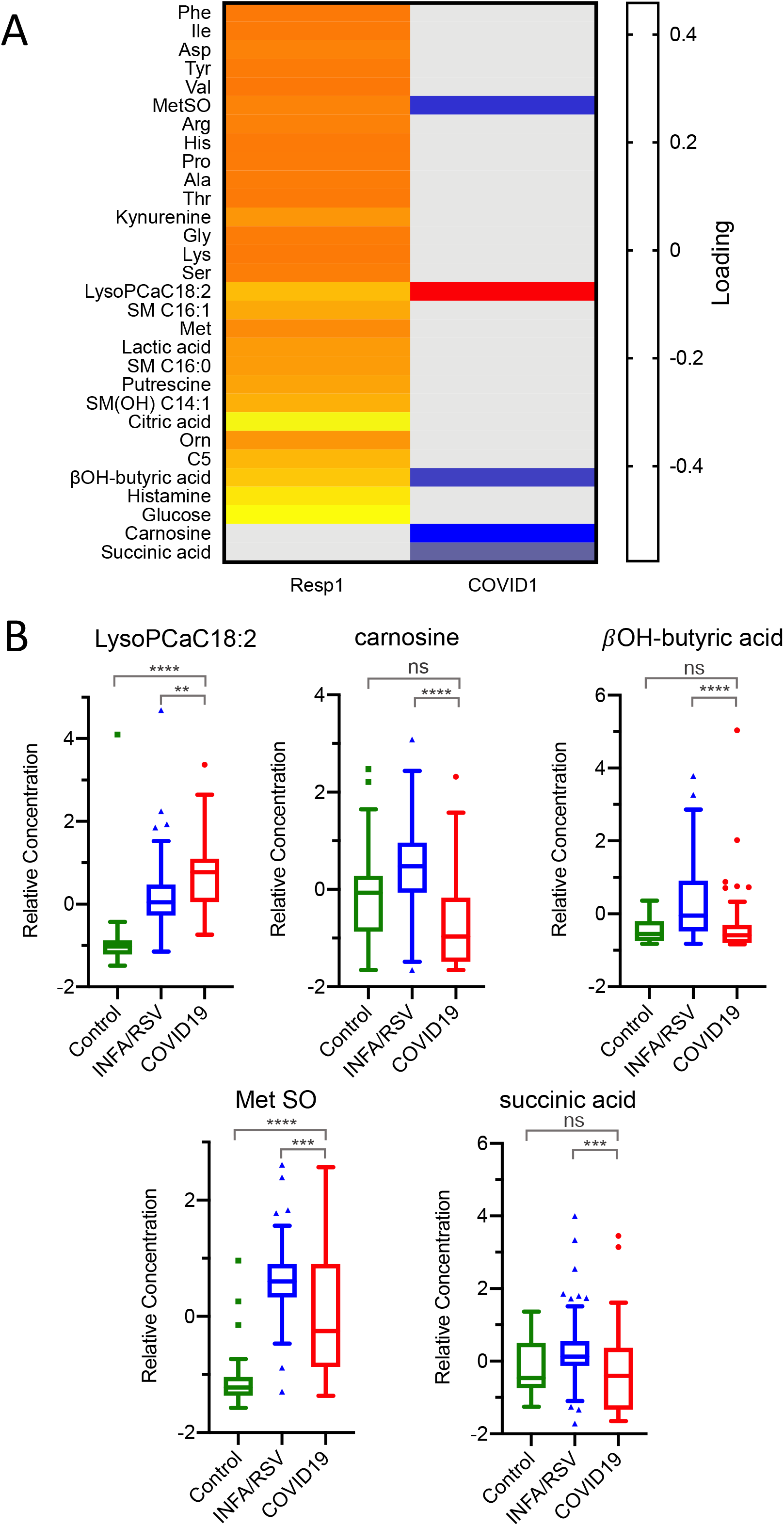
Feature selection from OPLS-DA models. (A) metabolite loadings for respiratory infection and COVID19 models highlighting the most significant features. For the respiratory infection model, the heatmap denotes the relative association of each metabolite with respect to asymptomatic controls. For the COVID19 model, the heatmap denotes the relative association of each metabolite with respect to influenza A/RSV patient group. (B) Boxplots showing relative concentrations of significant metabolites from the COVID19 model are mean-centered at zero. For metabolites presented in panel B, P<0.0001 by Kruskal-Wallis test. Significance of between-group means by post hoc Dunn’s test is given in each plot.

## Discussion

Using targeted LC-MS/MS-based metabolomics, we identified unique metabolite profiles associated with the nasopharynx of patients with common respiratory infections. We observed striking differences in signatures that could be used to differentiate asymptomatic controls from patients with COVID19, INFA or RSV. Furthermore, we identified a COVID-19-specific signature that was characterized by altered concentrations of LysoPCaC18:2, beta-hydroxybutyric acid, Met SO, succinic acid, and carnosine, relative to INFA and RSV.

While several metabolomics studies related to COVID-19 have emerged, the use of both targeted and untargeted approaches applied to a range of biosamples makes comparing results among studies challenging (4). The current study is unique as it employed a targeted approach to profile VTM acquired from standard-of-care swab kits from a diverse cohort of patients with qRT-PCR-confirmed COVID-19, IFNA or RSV, as well as asymptomatic controls. Although two previous studies have analyzed NP swabs, one study assessed VTM using matrix-assisted laser desorption/ionization mass spectrometry (MALDI-TOF MS)(5). The other study analyzed fresh swabs directly by ambient ionization methods including DESI and LD-REIMS and focused on lipid profiling (6). Both studies revealed diagnostic accuracies of >80%. At least three studies investigating the serum metabolome of COVID-19 patients identified changes in the tryptophan-kynurenine pathway associated with regulation of inflammation (7). We also observed an increase in kynurenine concentration in our respiratory model, but this metabolite was not COVID-19 specific. Our results are consistent with Bo Shen *et al*. (8) who also observed increased kynurenine concentration in non-COVID-19 patients presenting with symptoms of respiratory infection. Amino acids are decreased in interleukin-6 stratified COVID-19 patients compared to controls purportedly due to renal dysfunctional and marked alterations in nitrogen metabolism (7). In contrast, we saw an increase in amino acids in respiratory virus infection compared to control, yet this may reflect a difference in systemic (serum) vs local (nasopharynx) compartments sampled. Of the three studies involving serum metabolome analysis, Blasco *et al*. (9) calculated a diagnostic accuracy for COVID-19 as 74%.

A brief literature review of analytes in the COVID-19 metabolomic profile yields potential insight into the infection pathway (Fig 3). In particular, carnosine and LYSOC18:2 had strong loadings in the OPLS-DA model, with the former decreasing, and the latter increasing, in COVID-19 patients relative to patients with INFA or RSV. Carnosine, a naturally occurring dipeptide, has a wide range of protective effects in humans, which are largely attributed to its powerful antioxidant actions (10). Several mechanisms could explain the depleted levels of carnosine in COVID-19 patients. First, decreased carnosine levels may signify decreased production of the dipeptide by the host. The olfactory system is among the richest sources of carnosine in humans (11, 12), and carnosine present in the nasal swabs likely originated from the olfactory epithelium at the roof of the nasal cavity. The downregulation of carnosine could reflect decreased biosynthesis/secretion by olfactory sensory nerves or progressive loss of these neurons. Second, reduced carnosine levels may be the result of increased dipeptide degradation. Carnosine is largely metabolized by carnosinase-1 (CN1)(10). Although CN1 is expressed by the human olfactory epithelium (13), the nasal cavity is not typically considered a site of high carnosinase activity, such that intranasal administration of carnosine has been employed in a preclinical model of Parkinson disease as a means to avoid degradation by carnosinase (14). A third, and more probable explanation, is that the diminished carnosine levels indicate depleted carnosine stores. Our data show that even basal levels of the dipeptide are completely exhausted in COVID-19 patients. Saadah *et al*. (15) predicted that COVID-19-induced oxidative stress would result in carnosine depletion. The protective effects of carnosine are widely attributed to its antioxidant, anti-glycation, and anti-inflammatory properties (10). For instance, reduced circulating levels of low-density lipoprotein (LDL) has been associated with increased risk of acute kidney injury in COVID-19 patients (16), and carnosine, known to block lipid peroxynitrite-mediated modification of human LDL at physiological levels (17), may protect against LDL degradation. Intriguingly, recent papers suggest that carnosine may also protect against SARS-CoV-2 infection through more specific mechanisms. Molecular docking and modelling studies identified carnosine as the most promising drug candidate to prevent the binding of SARS-CoV-2 to the ACE2 receptor(15). Sustentacular cells of the olfactory system co-express the ACE2 receptor as well as TMPRSS2, a protease that facilitates viral entry, making these cells highly susceptible to SARS-CoV-2 (18). Infection of these cells has been implicated in anosmia9, a recognized symptom of COVID-19 (19). Given that the olfactory epithelium is a major producer of carnosine and this dipeptide’s vital neuroprotective role in this system (20), loss of carnosine may lead to olfactory nerve damage resulting in anosmia in COVID-19.

**Figure 3:**
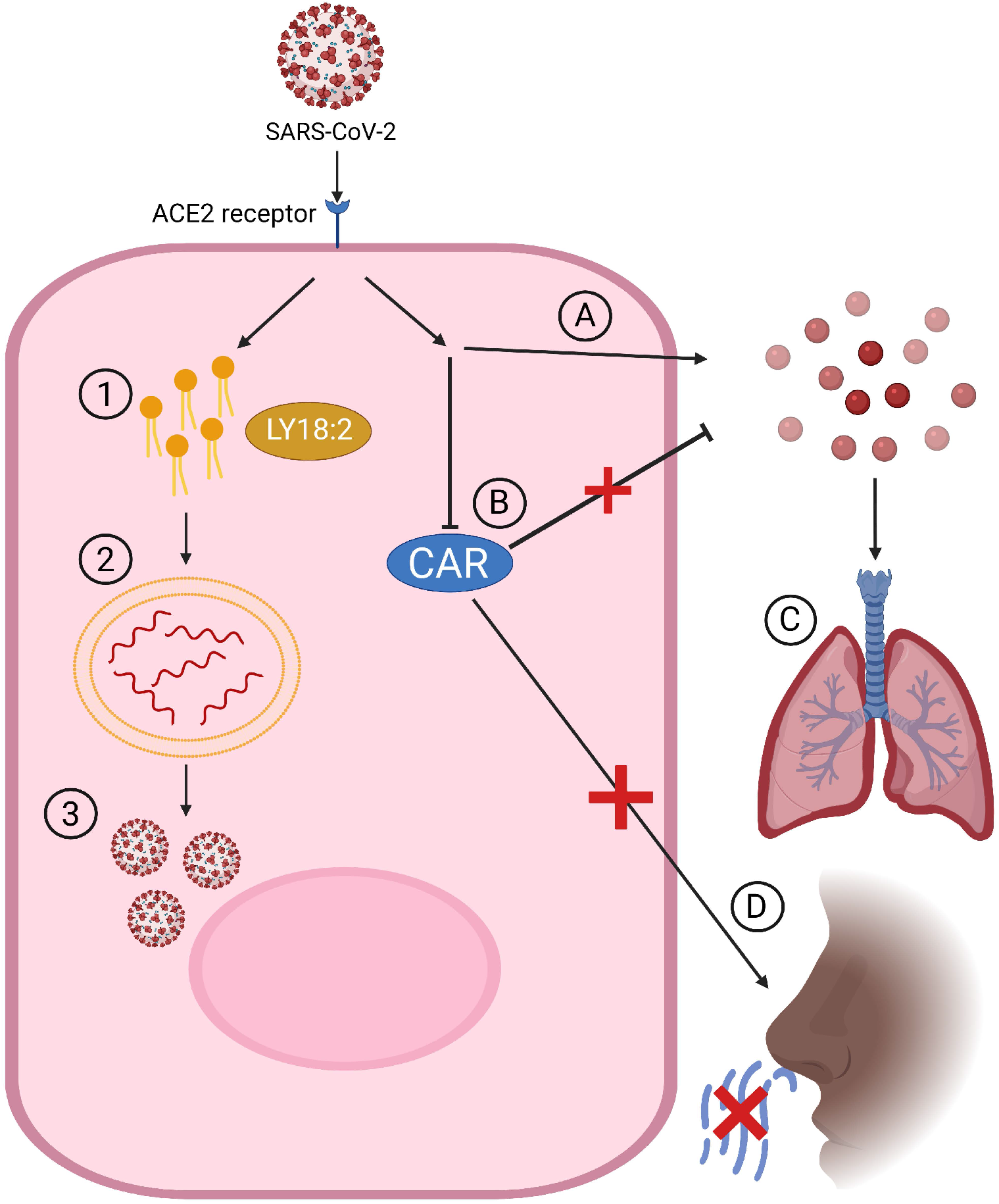
Schematic view of SARS-CoV-2 infection and potential mechanisms of symptom generation involving significantly altered metabolites. **(1)** After viral entry into the cell an increase in lipid generation occurs through viral hijacking of cell machinery. **(2)** Lipids are used to generate double membrane vesicles for replication. **(3)** Release of new Coronavirus. **(A)** SARS-CoV-2 leads to an elevation in oxidative stress by generation of reactive oxygen species (ROS). **(B)** Decreased levels of Carnosine results in a reduced ability for antioxidant clearance of ROS. **(C)** Oxidative stress and inflammation can damage the lungs and lead to further symptoms of COVID-19. **(D)** Reduction of Carnosine within the cells of the olfactory may be implicated in anosmia, a common symptom of COVID-19.

Severe cases of COVID-19 are associated with multi-organ damage arising from oxidative stress (21), raising the possibility of global depletion of carnosine in these patients and underscoring its therapeutic potential (22). Recently, Kulikova and colleagues (23) developed an analogue of carnosine, salicyl-carnosine, designed to circumvent the rapid degradation by serum carosinase. Salicyl-carnosine effectively mitigates the three prominent pathogenic hallmarks of COVID-19—oxidative stress, thrombosis, and inflammation—and has been proposed as a promising means to treat severe cases of COVID-19 (24).

Lysophosphatidylcholine (LysoPCaC18:2) is an endogenous bioactive phospholipid suggested to be important in coronavirus infection. The hijacking of host cells by the virus to create a proper environment for replication involves the generation of specialized vesicles of which LysoPCaC18:2 is required (25). Our study identified significantly elevated levels of LysoPCaC18:2 in COVID19 positive samples compared to INFA and RSV. In COVID19 viral infection, a change towards an increase in lipid generation has been demonstrated and further supports our finding (26, 27) especially whereby LysoPCaC18:2 was higher in COVID19 samples compared to healthy controls (28). Additionally, a study inhibiting Cytosolic Phospholipase A2α (cPLA2α) which produces lysophospholipids, had significant effects on lowering coronavirus RNA and protein accumulation due to the importance of phospholipids in the creation of replicative organelles, emphasizing the therapeutic potential of lipid metabolism pathways. Moreover, the same study found that inhibition of cPLA2α had no impact on the replication of influenza A thus, consistent with the COVID19-specific pattern of LysoPCaC18:2 concentration observed (29).

A limitation of our study was that extensive validation of the TMIC Prime kit for use with VTM was not conducted, including evaluation of matrix effects arising from VTM. While recovery of most synthetic metabolites was demonstrated at a single level in VTM, accuracy may have been affected by matrix interferences. For the purposes of this pilot study, we used the TMIC Prime kit as a rapid screening method to evaluate broad metabolite profiles in patients with respiratory diseases that will form the basis of more refined assays for individual metabolites in future studies. While internal standards corrected for recovery of analytes from VTM, we could not account for differences in yields of NP swabs. Furthermore, assay performance metrics for certain lipids such as LysoPCaC18:2 were not determined as we lacked a synthetic standard, and the internal standard used for this lipid was non-specific.

In conclusion, we demonstrated that the metabolome of the nasopharynx can be measured from clinical nasal swabs, and that metabolite profiles identified using machine learning methods can differentiate patients with COVID-19 from other respiratory virus infections (e.g. INFA/RSV). Our study identified key metabolites specifically altered in COVID19 such as carnosine and LysoPCaC18:2 that have previously been implicated in viral replication and symptom generation. This enables us to propose mechanisms contributing to viral infection and propagation as well as potential targets for COVID-19 therapy.

## Materials and Methods

### Ethics

All experimental protocols were approved by and conducted in accordance with the Queen’s University Health Sciences and Affiliated Teaching Hospitals Research Ethics Board (HSREB Files 6029794, 6029811).

### Sample Collection and qRT-PCR Analysis

Nasopharyngeal (NP) swabs were collected and stored in viral transport media (Copan Diagnostics, USA) from individuals being tested for SARS-CoV-2, INFA, and RSV at Kingston Health Sciences Centre (KHSC) and surrounding hospitals. Asymptomatic participants were recruited and tested for SARS-CoV-2 as part of a surveillance study testing medical and nursing students during the 2020 SARS-CoV-2 lockdown in Canada. Extraction of total RNA from VTM was performed using the Maxwell RSC Whole blood RNA/DNA kit (Promega Corporation, Madison, WI) on the Maxwell RSC 16 automated nucleic acid extractor in the KHSC clinical laboratory. The samples were tested for the presence of SARS-CoV-2 using laboratory-developed multiplex quantitative real-time PCR (qRT-PCR) assays. Briefly the qRT-PCR assay for SARS-CoV-2 targeting the Envelope (E) and RNA dependent RNA polymerase (RdRp) genes as previously described (Corman 2020). Samples for INFA, and RSV were tested using a clinically validated laboratory-developed multiplex qRT-PCR assay for INFA (matrix), and RSV (nucleoprotein).

Nasopharyngeal (NP) swabs positive for INFA and RSV were collected in 2019 - 2020, prior to the detection of SARS-CoV-2 in Canada, and stored at -80°C. Biological samples and demographic data were collected from patients within the circle of care. Samples were anonymized and de-identified so researchers were blind to the identity of the patients. Only secondary non-identifying data including age, biological sex, and travel history were provided. Under Ontario’s Personal Health Information Protection Act, all patients have the right to withhold or withdraw their consent for the use, access or disclosure of their Personal Health Information and patients are not disadvantaged if they refuse to participate. The requirement for written informed consent was waived by the Research Ethics Board since we used anonymized and de-identified samples provided for clinical testing.

## Metabolomic Analysis

### Sample Preparation and LC-MS/MS Analysis

Metabolite profiling kits (TMIC Prime) were acquired from The Metabolomics Innovation Centre (TMIC, Edmonton AB, Canada)(30, 31). The kit is capable of quantifying 141 analytes over six chemical classes using a combination of LC-MS/MS and flow-injection analysis (FIA)-MS/MS. Target analytes comprised organic acids, amino acids, biogenic amines, total hexoses, acylcarnitines and lipids (phosphatidylcholines (PC), lysophosphatidylcholines (LysoPC), sphingomyelins (SM), and hydroxy-sphingomyelins SM(OH)) (Supplementary Table 1). LC-MS/MS analysis was conducted using an ExionLC™ AC Series ultra-high-performance liquid chromatography system QTRAP® 5500 mass spectrometer (Sciex Canada, Concord, ON, Canada) in electrospray ionization (ESI) mode using optimized settings and MRM transitions provided by the manufacturer of the assay kit. For water-soluble analytes, separations were conducted on an Eclipse XDB-C18 HPLC column (3.5um, 3.0×100mm; Agilent, CA, USA) protected by a standard guard cartridge system (SecurityGuard™ Phenomenex®, CA, USA). For FIA of lipid-soluble analytes and glucose, samples were injected directly into the mass spectrometer via PEEK tubing.

50 µL of VTM was supplemented with appropriate internal standards and treated with 150 µL of 50% ethanol, homogenized by vortex mixing and sonication, and pelleted by centrifugation. The supernatant was dried on an N2 evaporator and re-dissolved in 50% ethanol. To assay organic acids, samples were transferred to a deep-well 96-well plate. The following 3 solutions were added to each well for derivatization: 1) 25 µL of 250 mM 3-nitrophenylhydrazine prepared in 50% methanol 2) 25 µL of 150 mM 1-ethyl-3-(3-dimethylaminopropyl) carbolimide prepared in methanol 3) 25 µL of 7.5% pyridine prepared in 75% methanol. The plate was shaken at room temperature for 2 h. 375 µL of water was added, and the plate was shaken for 20 minutes at room temperature. 125 µL was transferred to a new plate and diluted with 375 µL of 50% methanol. For LC separation, 10 µL of the sample was injected into the LC-MS/MS system in negative ionization mode, using a flow rate of 300 µL/min where mobile phase A consisted of 0.01% formic acid (FA) in water, and mobile phase B consisted of 0.01% FA in methanol. The linear gradient elution profile for mobile phase B was: t=0 min, 30%; t=1.5 min, 30%; t=12.5 min, 85%; t=12.51 min, 100%.

For assay of amino acids, biogenic amines, and lipids, a second aliquot of VTM were prepared as described above. Samples were aliquoted onto a filter paper disc in each well of the 96-well filter plate. Samples were dried for 30 minutes on an N2 evaporator. For derivatization of amino acids and biogenic amines, a 5% solution of phenyl-isothiocyanate (PITC) was prepared in equal parts ethanol/pyridine/water. 50 µL of the 5% PITC solution was added to each filter paper. The plate was covered and incubated at room temperature for 20 minutes. The plate was dried for 90 min to remove excess liquid. 300 µL of 5 mM ammonium acetate in methanol was added to each well, and the plate was shaken at room temperature for 30 minutes to extract analytes from the filter paper. The extract was collected by centrifugation. For LC-MS/MS analysis of derivatized amino acids and biogenic amines, 62.5 µL of the extract was combined with 62.5 µL of water in a new 96-well plate and shaken. 10 µL of the sample was injected into the LC-MS/MS system in positive ionization mode using a flow rate of 500 µL/min, where mobile phase A consisted of 0.2% FA in water, and mobile phase B consisted of 0.2% FA in acetonitrile. The linear gradient elution profile for mobile phase B was: t=0 min, 0%; t=0.5 min, 0%; t=5.6 min, 95%; t=6.5 min, 95%. For FIA-MS/MS of underivatized glucose, acylcarnitines and lipids, 25 µL of the extract was combined with 125 µL of FIA buffer in a new plate. FIA buffer was prepared by adding 9 mL of 0.1% FA to 260 mL of methanol and was used as the sample diluent and mobile phase. 20 µL of the sample was injected for FIA analysis using the following flow rate profile: t=0 min, 30 µL/min; t=1.6 min, 30 µL/min; t=2.4 min, 200 µL/min; t=2.8, 200 µL/min. Two injections were conducted for FIA analysis: one in negative ionization mode for measurement of glucose, and a second injection in positive ionization mode for measurement of acylcarnitines and lipids.

Quantification of analytes measured by LC-MS/MS was based on isotope dilution and quadratic calibration lines for each analyte. Peak integration and analyte quantification were completed using Analyst 1.7 (Sciex). Analytes measured by FIA-MS/MS were quantified using a relative quantification approach using a single representative internal standard for each analyte class. Supplementary Table 2 presents assay performance parameters for prioritized metabolites measured by LC-MS/MS. Four quality control samples based on solution standards (QC 1-3) and a low-level spiked VTM sample were measured 3 times on each of 3 assay days. 89% of measurements in solution standards were within 20% of target values, and all but one analyte exhibited total CVs of <20%. Total CVs for spiked VTM was more variable, with only 52% of analyte measurements exhibiting CVs of <20%. Mean % differences from target concentration was +28% (range: -14.3-112%). Poorer assay performance metrics in VTM are likely due to the presence of matrix interferences that we were unable to evaluate in the current study.

## Statistical Analysis

Our fully open and reproducible analysis pipeline, written in **R** (v 4.3), is available online (https://github.com/ColauttiLab/COVID-Metabolomics) and detailed in the Supplementary Methods. Briefly, we first scaled samples to baseline levels observed in VTM and then autoscaled each metabolite to a mean of zero and unit standard deviation. We used individual univariate models to test whether a metabolite differed among patients from the four different categories. We then imputed missing values (N = 87 of 7,735) and randomly divided our data into a model-building test dataset (50% of data) and a model-testing validation dataset (50% of data) that was excluded from the model-building pipeline. Metabolites with significant differences among groups in the test dataset, after false-discovery rate adjustment, were included in multivariate partial least-squares discriminant analysis (PLS-DA) models as implemented by the *opls* function from the **ropls** package (32). The accuracy, sensitivity, and specificity of the multivariate models were tested on the validation dataset. Statistical analysis of individual metabolites among patient groups (i.e. univariate models) were performed using a non-parametric ANOVA (Kruskal-Wallis) with a post hoc Dunn’s test. Differences in metabolite concentrations were determined to be significant if P<0.05.

## Data Availability

Our fully open and reproducible analysis pipeline, written in R (v 4.3), is available online (https://github.com/ColauttiLab/COVID-Metabolomics) and detailed in the Supplementary Methods.

## Acknowledgments

The authors would like to thank KHSC and the Clinical Microbiology laboratory for providing banked nasopharyngeal swab specimens and access to required patient details. We thank TMIC for assistance with use of Prime kits.

## Author contributions

MK, SB, KT, PS, DR and SV designed the study and applied for grant funding. PS, HW and AKE provided nasopharyngeal specimens. MK and SB performed the LC-MS/MS experiments. MK, SB, CS and RC analyzed the data. All authors helped with the interpretation of the findings. RC and CS provided computational modelling expertise and results analysis. PS, MK, SB, CS and KT were involved in the writing of the initial manuscript drafts, with later input from the remaining authors.

