## Supplementary document for "Small-molecule metabolome identifies potential therapeutic targets against COVID-19"

NOTE: Raw data and fully reproducible code for this project are available on GitHub:
http://bit.ly/COVID-Metabolomics

### Contents

**Figure S1**: Experimental Workflow 2

**Table S1**: List of metabolites assessed using the TMIC Prime kit 3

**Figure S2**: Correlation of CTs with selected metabolites from the COVID model 6

**Table S2**: Assay performance parameters for prioritized analytes measured by LC-MS/MS 7

**Supplementary Methods** 8

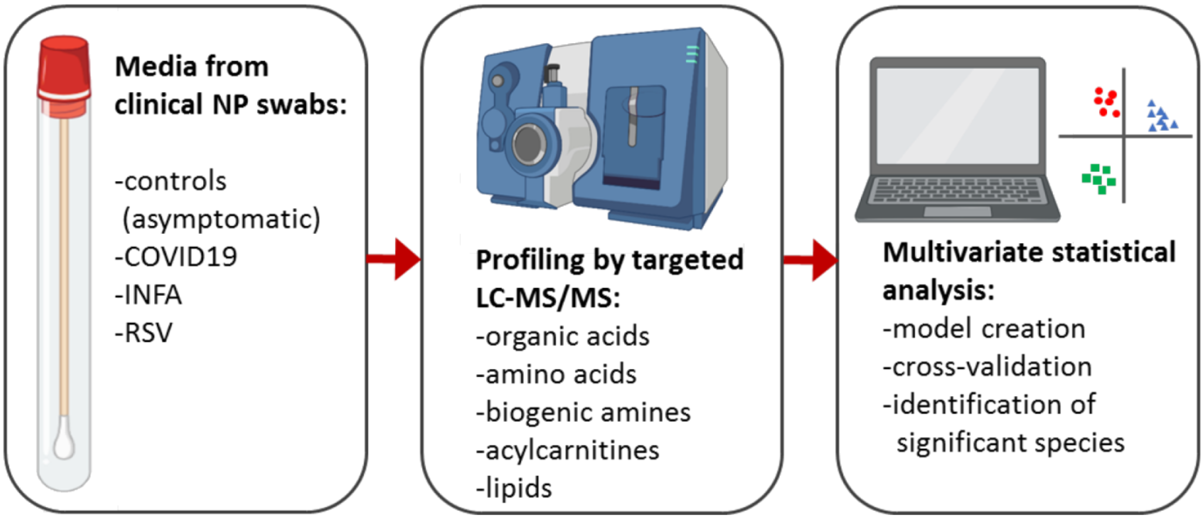


**Figure S1: Experimental workflow.** Viral transport medium from clinical nasopharyngeal swabs was analyzed using a TMIC Prime kit involving chemical derivatization, and liquid chromatography-tandem mass spectrometry. Multivariate and univariate statistical analyses were conducted to identify significant features, which we attempt to rationalize in the context of the pathogenesis of viral infection.

**Supplementary Table 1: List of metabolites assessed using the TMIC Prime kit.**

| Acylcarnitines | | | |
|---|---|---|---|
| C0 | Carnitine | C10:1 | Decenoylcarnitine |
| C2 | Acetylcarnitine | C10 | Decanoylcarnitine |
| C3:1 | Propenoylcarnitine | C12:1 | Dodecenoylcarnitine |
| C3 | Propionylcarnitine | C12 | Dodecanoylcarnitine |
| C4:1 | Butenylcarnitine | C14:2 | Tetradecadienylcarnitine |
| C4 | Butyrylcarnitine | C14:1* | Tetradecenoylcarnitine |
| C3OH | Hydroxypropionylcarnitine | C14 | Tetradecanoylcarnitine |
| C5:1 | Tiglylcarnitine | C12DC | Dodecanedioylcarnitine |
| **C5** | **Valerylcarnitine** | C14:2OH | Hydroxytetradecadienylcarnitine |
| C4OH | Hydroxybutyrylcarnitine | C14:1OH | Hydroxytetradecenoylcarnitine |
| C6:1 | Hexenoylcarnitine | C16:2 | Hexadecadienylcarnitine |
| C6 | Hexanoylcarnitine | C16:1 | Hexadecenoylcarnitine |
| C5OH | Hydroxyvalerylcarnitine | C16 | Hexadecanoylcarnitine |
| C5:1DC | Glutaconylcarnitin | C16:2OH | Hydroxyhexadecadienylcarnitine |
| C5DC | Glutarylcarnitine | C16:1OH | Hydroxyhexadecenoylcarnitine |
| C8 | Octanoylcarnitine | C16OH | Hydroxyhexadecanoylcarnitine |
| C5MDC | Methylglutarylcarnitine | C18:2 | Octadecadienylcarnitine |
| C9 | Nonaylcarnitine | C18:1 | Octadecenoylcarnitine |
| C7DC | Pimelylcarnitine | C18 | Octadecanoylcarnitine |
| C10:2 | Decadienylcarnitine | C18:1OH | Hydroxyoctadecenoylcarnitine |
| **Amino Acids** | | | |
| **Ala*** | **Alanine** | **Lys*** | **Lysine** |
| **Arg*** | **Arginine** | **Met*** | **Methionine** |
| Asn* | Asparagine | **Orn*** | **Ornithine** |
| **Asp*** | **Aspartate** | **Phe*** | **Phenylalanine** |
| Cit* | Citrulline | **Pro*** | **Proline** |
| Gln* | Glutamine | **Ser*** | **Serine** |
| Glu | Glutamate | **Thr*** | **Threonine** |
| **Gly*** | **Glycine** | Trp | Tryptophan |
| **His*** | **Histidine** | **Tyr*** | **Tyrosine** |
| **Ile*** | **Isoleucine** | **Val*** | **Valine** |
| Leu* | Leucine | | |
| **Benzenoids** | | | |
| Tyramine | | Homovanillic acid | |
| Hippuric acid | | | |

| | | | |
|---|---|---|---|
| **Biogenic Amines** | | | |
| Ac-Orn | Acetylornithine | **Met-SO*** | **Methionine sulfoxide** |
| ADMA | Asymmetric dimethylarginine | PEA | Phenylethylamine |
| alpha-AAA | alpha-Aminoadipic acid | Putrescine* | Putrescine |
| c4-OH-Pro | cis-4-Hydroxyproline | Sarcosine* | Sarcosine |
| **Carnosine** | **Carnosine** | Serotonin | Serotonin |
| Creatinine* | Creatinine | Spermidine* | Spermidine |
| Dopamine | Dopamine | Spermine* | Spermine |
| **Histamine*** | **Histamine** | t4-OH-Pro* | trans-4-Hydroxyproline |
| **Kynurenine*** | **Kynurenine** | Taurine* | Taurine |
| Methylhistidine | Methylhistidine | total DMA* | Dimethylamine |
| **Glycerophospholipids** | | | |
| LysoPC a C14:0* | LysoPhosphatidylcholine acyl C14:0 | LysoPC a C28:1 | LysoPhosphatidylcholine acyl C28:1 |
| LysoPC a C16:1* | LysoPhosphatidylcholine acyl C16:1 | LysoPC a C28:0 | LysoPhosphatidylcholine acyl C28:0 |
| LysoPC a C16:0* | LysoPhosphatidylcholine acyl C16:0 | PC aa C32:2 | Phosphatidylcholine diacyl C32:2 |
| LysoPC a C17:0* | LysoPhosphatidylcholine acyl C17:0 | PC ae C36:0 | Phosphatidylcholine acyl-alkyl C36:0 |
| LysoPC a C18:2* | LysoPhosphatidylcholine acyl C18:2 | PC aa C36:6 | Phosphatidylcholine diacyl C36:6 |
| LysoPC a C18:1* | LysoPhosphatidylcholine acyl C18:1 | PC aa C36:0 | Phosphatidylcholine diacyl C36:0 |
| LysoPC a C18:0* | LysoPhosphatidylcholine acyl C18:0 | PC aa C38:6 | Phosphatidylcholine diacyl C38:6 |
| LysoPC a C20:4 | LysoPhosphatidylcholine acyl C20:4 | Pc aa C38:0 | Phosphatidylcholine diacyl C38:0 |
| LysoPC a C20:3 | LysoPhosphatidylcholine acyl C20:3 | Pc ae C40:6 | Phosphatidylcholine acyl-alkyl C40:6 |
| LysoPC a C24:0 | LysoPhosphatidylcholine acyl C24:0 | PC aa C40:6 | Phosphatidylcholine diacyl C40:6 |
| LysoPC a C26:1 | LysoPhosphatidylcholine acyl C26:1 | PC aa C40:2 | Phosphatidylcholine diacyl C40:2 |
| LysoPC a C26:0 | LysoPhosphatidylcholine acyl C26:0 | PC aa C40:1 | Phosphatidylcholine diacyl C40:1 |
| **Lipids and lipid-like molecules** | | | |
| Butyric acid | | | |
| **Organic acids and derivatives** | | | |
| Betaine | | **Citric acid** | |
| Creatine | | **Succinic acid** | |
| Diacetylspermine | | Fumaric acid* | |
| **Lactic acid*** | | Pyruvic acid | |
| **beta-Hydroxybutyric acid*** | | Isobutyric acid | |
| alpha-Ketoglutaric acid | | Methylmalonic acid | |
| DOPA | l-3,4-dihydroxyphenylalanine | Nitro-Tyr | Nitrotyrasine |
| Propionic acid | | p-Hydroxyhippuric acid | para-hydroxyhippuric acid |
| **Organoheterocyclic compounds** | | | |
| Indole acetic acid | | Uric acid* | |
| **Organic nitrogen compounds** | | | |
| Choline* | Choline | TMAO | Trimethylamine N-oxide |
| **Hexose** | | | |
| **Glucose** | | | |
| **Phenylpropanoids and polyketides** | | | |
| HPHPA | 3-(3-Hydroxyphenyl)-3-hydroxypropanoic acid | | |

| Sphingomyelins | | | |
|---|---|---|---|
| SM(OH) C14:1 | Hydroxysphingomyeline C14:1 | SM C18:0 | Sphingomyeline C18:0 |
| **SM C16:1** | **Sphingomyeline C16:1** | SM C20:2 | Sphingomyeline C20:2 |
| **SM C16:0*** | **Sphingomyeline C16:0** | SM(OH) C22:2 | Hydroxysphingomyeline C22:2 |
| SM(OH) C16:1 | Hydroxysphingomyeline C16:1 | SM(OH) C22:1 | Hydroxysphingomyeline C22:1 |
| SM C18:1 | Sphingomyeline C18:1 | SM(OH) C24:1 | Hydroxysphingomyeline C24:1 |

*Significant metabolites P=<0.05; Red colouring: Metabolite measured as two times above VTM.

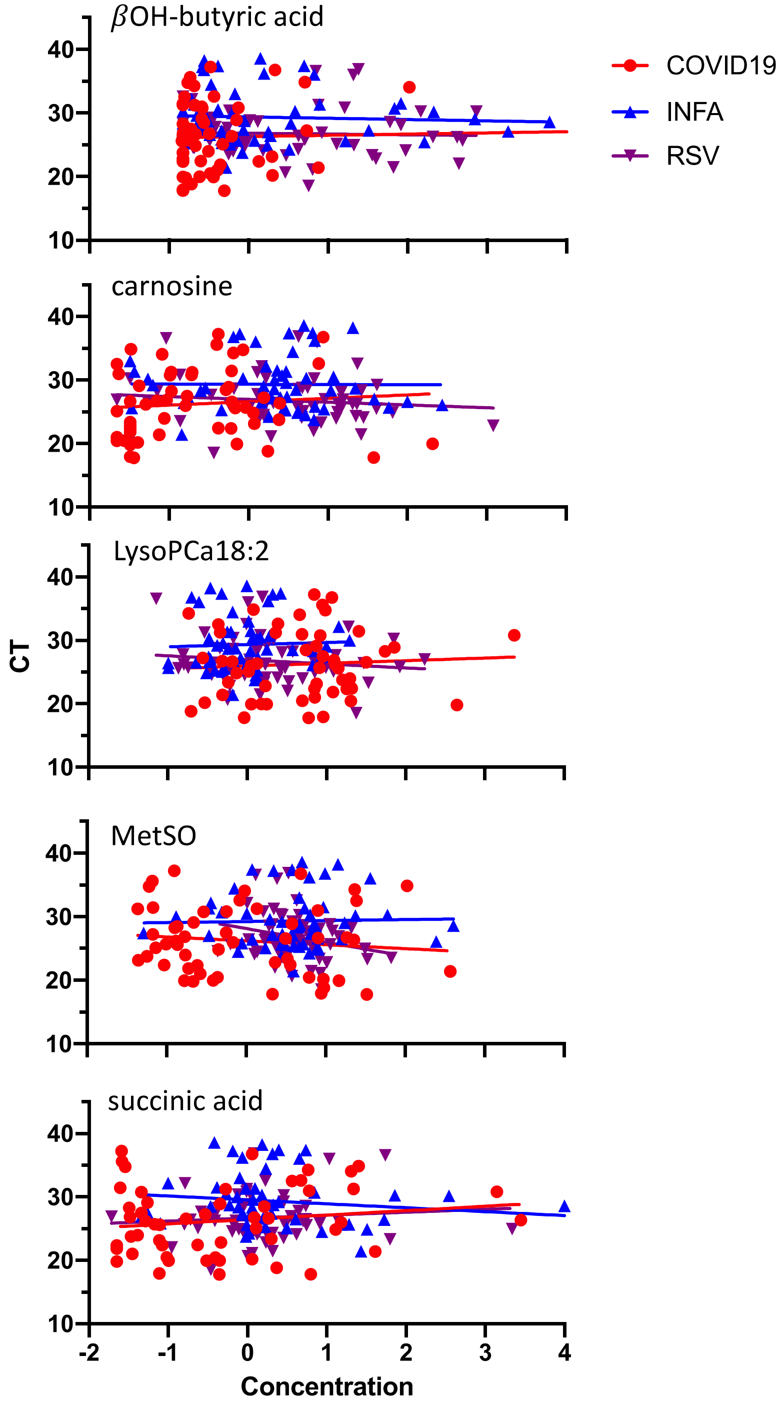


**Figure S2: Correlation of cycle threshold (CT) with selected metabolites from the COVID model.**

#### Supplementary Table 2: Assay performance parameters for prioritized analytes measured by LC-MS/MS

| Analyte ID | QC1[1] | | | | QC2[1] | | | | QC3[1] | | | | VTM[1,4] | | | | |
|---|---|---|---|---|---|---|---|---|---|---|---|---|---|---|---|---|---|
| | [Target] µM | [Mean] µM | Diff.[2] % | CV[3] % | [Target] µM | [Mean] µM | Diff. [2] % | CV[3] % | [Target] µM | [Mean] µM | Diff. [2] % | CV[3] % | FC[4] | [Target] µM | [Mean] µM | Diff. [2] % | CV[3] % |
| Ala | 160 | 203.1 | -26.9 | 11.0 | 640 | 764.0 | -19.4 | 10.7 | 1280 | 1438.9 | -12.4 | 10.7 | 28.7 | 40 | 54.0 | 35.0 | 20.0 |
| Arg | 40 | 46.5 | -16.3 | 4.8 | 160 | 176.7 | -10.4 | 6.2 | 320 | 355.9 | -11.2 | 4.7 | 18.1 | 10 | 13.5 | 35.2 | 8.5 |
| βOH-butyric acid | 12.5 | 13.5 | -8.0 | 5.6 | 50 | 51.7 | -3.5 | 5.6 | 100 | 104.7 | -4.7 | 4.4 | 6.8 | 10 | 8.6 | -14.3 | 5.4 |
| carnosine | 8 | 9.4 | -17.3 | 9.0 | 32 | 35.7 | -11.7 | 7.8 | 64 | 71.6 | -11.9 | 5.5 | 1.2 | 2 | 3.2 | 58.0 | 7.5 |
| citric acid | 12.5 | 13.4 | -6.9 | 9.3 | 50 | 49.5 | 1.0 | 5.8 | 100 | 99.9 | 0.1 | 3 | 0.4 | 10 | 11.6 | 16.0 | 3.5 |
| Gly | 200 | 238.6 | -19.3 | 16.5 | 800 | 829.9 | -3.7 | 6.1 | 1600 | 1647.8 | -3.0 | 10.9 | 14.0 | 50 | 51.6 | 3.2 | 33.0 |
| His | 40 | 34.6 | 13.4 | 6.8 | 160 | 134.1 | 16.2 | 6.8 | 320 | 265.4 | 17.0 | 7.3 | 25.3 | 10 | 13.1 | 31.2 | 50.1 |
| histamine | 8 | 8.4 | -4.6 | 14.4 | 32 | 33.2 | -3.8 | 11.1 | 64 | 65.4 | -2.2 | 14.8 | 4.9 | 2 | 3.4 | 71.0 | 15.8 |
| Ile | 40 | 45.0 | -12.5 | 6.2 | 160 | 159.2 | 0.5 | 8.1 | 320 | 306.1 | 4.3 | 10.3 | 26.0 | 10 | 13.0 | 29.8 | 28.7 |
| kynurenine | 40 | 50.8 | -27.1 | 6.6 | 160 | 178.8 | -11.7 | 5.0 | 320 | 348.1 | -8.8 | 9.9 | 7.8 | 10 | 8.3 | -17.0 | 10.6 |
| lactic acid | 125 | 137.0 | -9.6 | 7.5 | 500 | 517.1 | -3.4 | 5.1 | 1000 | 1002.0 | -0.2 | 4.7 | 4.4 | 100 | 115.0 | 15.0 | 7.7 |
| Lys | 40 | 51.3 | -28.2 | 11.4 | 160 | 183.2 | -14.5 | 8.7 | 320 | 328.2 | -2.6 | 9.7 | 24.3 | 10 | 13.6 | 35.8 | 14.6 |
| Met | 40 | 44.5 | -11.3 | 9.7 | 160 | 173.0 | -8.1 | 4.0 | 320 | 335.6 | -4.9 | 7.5 | 22.8 | 10 | 11.9 | 19.0 | 7.9 |
| MetSO | 8 | 8.5 | -6.8 | 17.4 | 32 | 33.5 | -4.8 | 8.6 | 64 | 72.1 | -12.7 | 8.2 | 28.5 | 2 | 2.3 | 16.0 | 23.6 |
| Orn | 8 | 9.6 | -20.0 | 9.2 | 32 | 33.5 | -4.8 | 10.8 | 64 | 62.4 | 2.5 | 13.6 | 8.1 | 2 | 2.9 | 45.0 | 85.0 |
| Phe | 40 | 48.2 | -20.6 | 8.4 | 160 | 184.7 | -15.4 | 6.5 | 320 | 342.6 | -7.0 | 5.5 | 23.8 | 10 | 13.3 | 33.4 | 24.4 |
| Pro | 80 | 98.3 | -22.9 | 12.8 | 320 | 373.6 | -16.7 | 4.9 | 640 | 652.4 | -1.9 | 8.0 | 21.6 | 20 | 24.4 | 22.0 | 18.2 |
| putrescine | 0.8 | 1.2 | -45.1 | 58.2 | 3.2 | 3.7 | -17.0 | 21.6 | 6.4 | 6.5 | -1.5 | 11.2 | 7.7 | 0 | 0.3 | 70.0 | 29.9 |
| Ser | 40 | 44.3 | -10.8 | 17.7 | 160 | 165.2 | -3.3 | 13.7 | 320 | 364.1 | -13.8 | 16.1 | 26.6 | 10 | 21.2 | 112.0 | 137.7 |
| succinic acid | 2.5 | 2.5 | 3.8 | 0.5 | 10 | 9.6 | 3.7 | 3.1 | 20 | 19.3 | 3.6 | 1.5 | 1.5 | 2 | 2.3 | 15.6 | 12.9 |
| Thr | 40 | 48.8 | -22.1 | 12.1 | 160 | 172.1 | -7.6 | 8.3 | 320 | 313.4 | 2.0 | 9.6 | 18.7 | 10 | 12.5 | 25.2 | 58.1 |
| Tyr | 40 | 39.1 | 2.3 | 9.9 | 160 | 149.7 | 6.5 | 4.8 | 320 | 274.3 | 14.3 | 13.9 | 9.1 | 10 | 8.6 | -13.6 | 24.9 |
| Val | 80 | 93.0 | -16.3 | 9.8 | 320 | 350.9 | -9.7 | 6.9 | 640 | 702.8 | -9.8 | 7.8 | 34.7 | 20 | 24.6 | 23.0 | 27.9 |

[1] Quality control material based on solution standards (QC1-3), or spiked viral transport medium (VTM). Each QC sample was assayed 9 times over 3 days.

[2] % difference of mean measured concentration relative to target.

[3] Total coefficients of variation based on 9 measurements over 3 days.

[4] Fold-change of mean concentration in blank VTM (N=12) as compared with mean concentration in patient samples. All other parameters for VTM are based on spiked VTM samples.

### Supplementary Methods

**NOTE: Raw data and fully reproducible code for this project are available on GitHub: http://bit.ly/COVID-Metabolomics**

#### Setup

##### Basic setup for plotting and data handling

```
library(tidyverse) # Tools for data science (graphing, data reorganizing, etc.)
library(ropls)

# Some custom graphing stuff
source("./theme_pub.R")
theme_set(theme_pub())
```

##### User-parameters

```
flipResp<-T # If True, reverse main axis scaling for respiratory model
flipCOVID<-T # If True, reverse main axis scaling for COVID model
```

##### Load data

```
featDatA<-read.csv("./data/FeatDatA.csv") # Features selected from resDat using Subset A
featDatB<-read.csv("./data/FeatDatB.csv") # Features selected from resDat using Subset B
featDatC<-read.csv("./data/FeatDatC.csv") # Features selected from resDat using Subset C
featDatCT<-read.csv("./data/FeatDatCT.csv") # Features selected from resDat using Subset based on correlation with CT
```

#### Pipeline Details

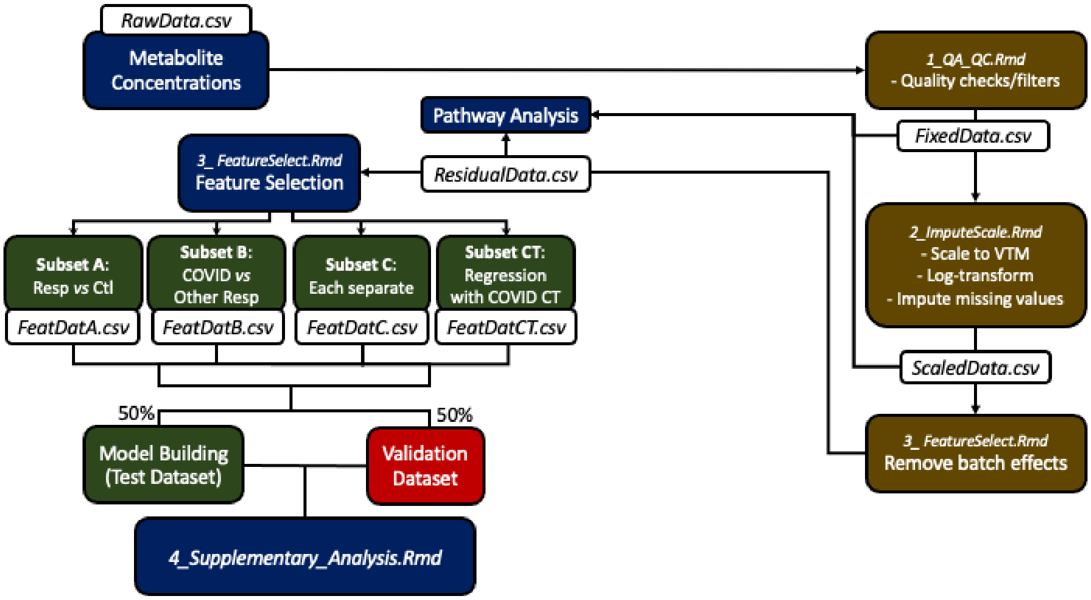


*Analysis Pipeline*

#### PLS-DA - all groups

Full Partial Least Squares Discriminant Analysis with all 4 groups (and 3 orthogonal predictor axes)

NOTE: This is used for graphing purposes only. For predictive models, see OPLS-DA models, below.

```
# RDA Model
# Setup for grid search with Leave-One-Out Cross-Validation (LOOC using the test-building
subset of data)
FULLdat<-featDatC %>% # Dataset with new encoding
  filter(Class.name %in% c("Control","COVID19","Influenza","RSV")) %>% # Remove VTM
  column_to_rownames("Sample.Name")

DescNames<-c("Batch.Number","Class.name","Sex","Age","CT","OrigClass") # Response Variabl
e
Concs<-names(FULLdat)[!names(FULLdat) %in% DescNames] # Predictor Variables


# Organize data for opls
metData<-FULLdat[,Concs] # Metabolite data
patClass<-FULLdat[,"Class.name"] # Predictors
# Set row.names
names(patClass)<-row.names(FULLdat)

# Model of full data for plotting
FULLmod<-opls(metData, patClass, predI=3, fig.pdfC="none")

## PLS-DA
## 210 samples x 31 variables and 1 response
## standard scaling of predictors and response(s)
##       R2X(cum) R2Y(cum) Q2(cum) RMSEE pre ort pR2Y  pQ2
## Total    0.663    0.395   0.334 0.343   3   0 0.05 0.05
```

NOTE: No confusion matrix is calculated here (no cross-validation). The purpose is to see whether samples form distinct groups, and factor loadings, rather than to generate and test predictions from the model (that is done below).

##### PLS-DA axis plots

```
pDatF<-as.data.frame(FULLmod@scoreMN)
pDatF$Class<-as.factor(FULLdat$Class.name)
pDatF$Age<-as.factor(FULLdat$Age)
pDatF$Sex<-as.factor(FULLdat$Sex)

ggplot(aes(x=p1,y=p2,group=Class,fill=Class,shape=Class),data=pDatF) +
  stat_ellipse(aes(colour=Class),size=1.2, alpha=0.8) +
  geom_point(size=3,alpha=0.8) +
  scale_fill_manual(values=c("#989788","#E54F6D","#008BF8","#623CEA","#E7EBC5")) +
  scale_colour_manual(values=c("#989788","#E54F6D","#008BF8","#623CEA")) +
  scale_shape_manual(values=c(22,21,24,25,22))
```

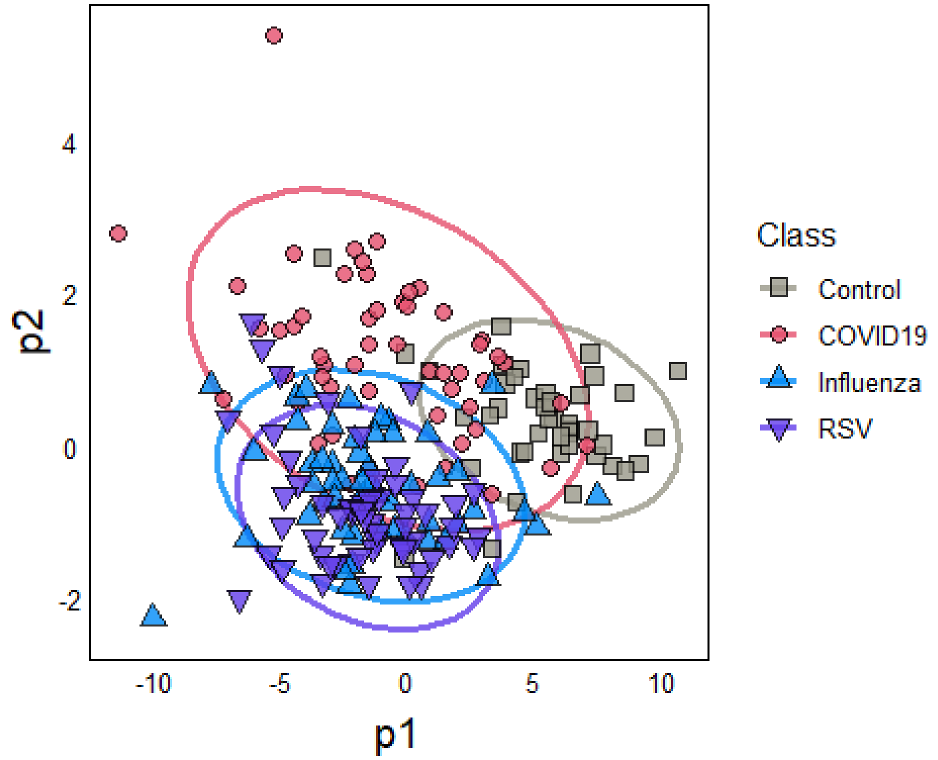


```r
ggplot(aes(x=p1,y=p3,group=Class,fill=Class,shape=Class),data=pDatF) +
  stat_ellipse(aes(colour=Class),size=1.2, alpha=0.8) +
  geom_point(size=3,alpha=0.8) +
  scale_fill_manual(values=c("#989788","#E54F6D","#008BF8","#623CEA","#E7EBC5")) +
  scale_colour_manual(values=c("#989788","#E54F6D","#008BF8","#623CEA")) +
  scale_shape_manual(values=c(22,21,24,25,22))
```

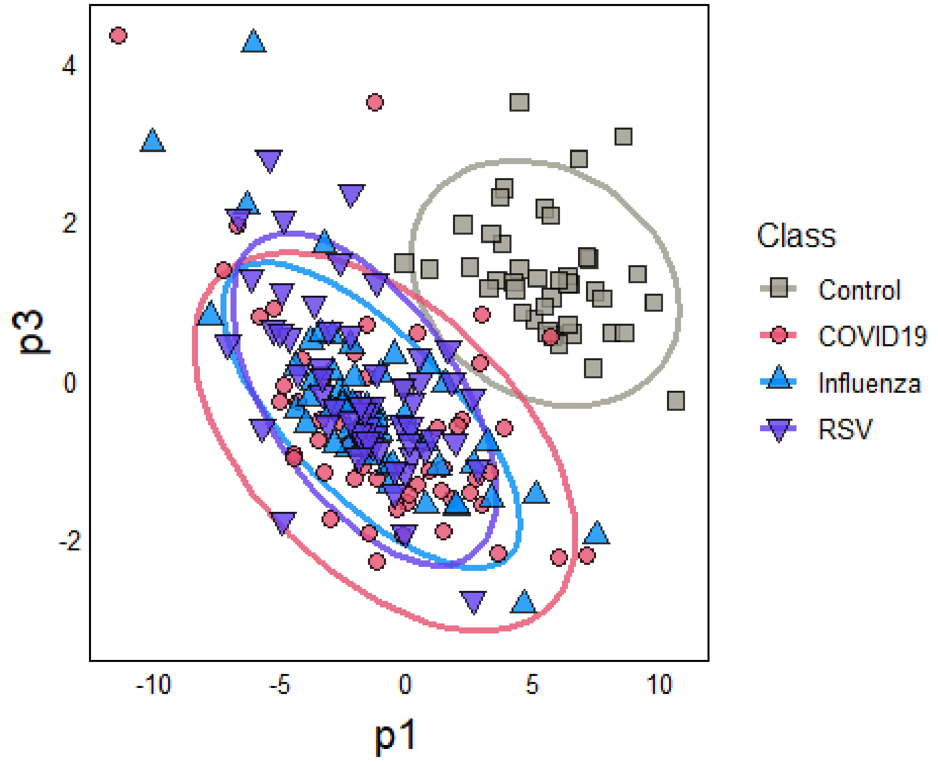

```
ggplot(aes(x=p2,y=p3,group=Class,fill=Class,shape=Class),data=pDatF) +
  stat_ellipse(aes(colour=Class),size=1.2, alpha=0.8) +
  geom_point(size=3,alpha=0.8) +
  scale_fill_manual(values=c("#989788","#E54F6D","#008BF8","#623CEA","#E7EBC5")) +
  scale_colour_manual(values=c("#989788","#E54F6D","#008BF8","#623CEA")) +
  scale_shape_manual(values=c(22,21,24,25,22))
```

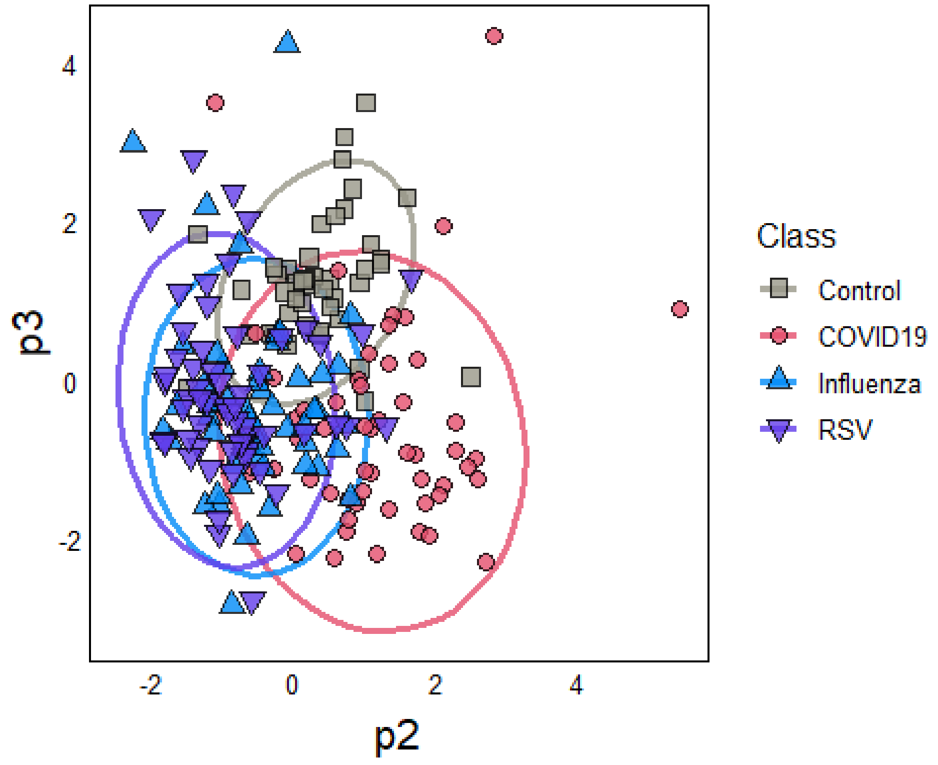


#### Export PLD-DA plotting data

```
#write.csv(pDatF,"./pDat/FULLdat.csv")
```

#### Graph of loadings:

```
Loadings<-as.data.frame(FULLmod@loadingMN)
Loadings$Metabolite<-row.names(Loadings)
heatDat<-gather(Loadings,Axis,Loading,all_of(names(Loadings[-4])))
heatDat<-as.data.frame(heatDat)

ggplot(aes(x=Axis,y=Metabolite,fill=Loading),data=heatDat) + geom_tile() +
  facet_grid(~ Axis, scales = "free_x", space = "free_x") +
  scale_fill_gradientn(colours=c("#008BF8","#E7EBC5","#E54F6D"))
```

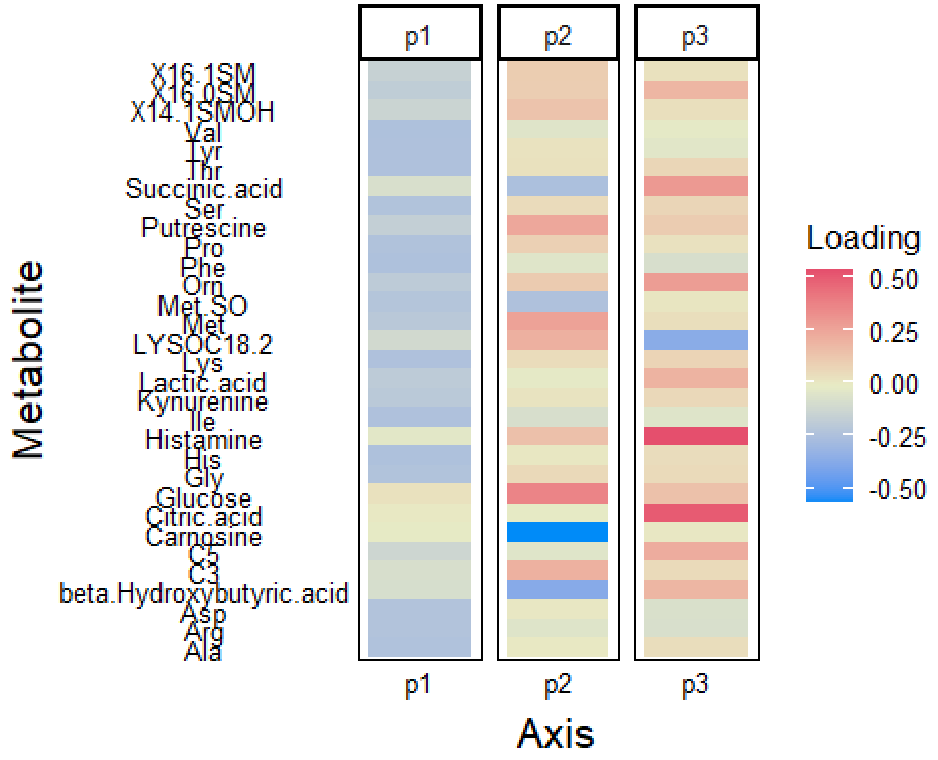


##### Export FULL model loadings data

```
#write.csv(heatDat,"./pDat/FULLload.csv")
```

#### OPLS-DA with FS

Orthogonal PLS used here for models based on two bins. NOTE: the x-axis is the orthogonal predictor, a second (y-axis) is added only for plotting purposes.

##### Control vs All respiratory

```
# Respiratory Only
RESPdat<-featDataA %>% # Dataset with new encoding
  filter(Class.name %in% c("Control","COVID19","Influenza","RSV")) %>%
  column_to_rownames("Sample.Name")
RESPdat$OrigClass<-RESPdat$Class.name
RESPdat$Class.name<-recode_factor(RESPdat$Class.name, COVID19 = "Resp",
                                   Influenza = "Resp", RSV = "Resp")

DescNames<-c("Batch.Number","Class.name","Sex","Age","CT","OrigClass") # Response Variabl
e
Concs<-names(RESPdat)[!names(RESPdat) %in% DescNames] # Predictor Variables

# Organize data for opls
metData<-RESPdat[,Concs] # Metabolite data
patClass<-RESPdat[,"Class.name"] # Predictors
# Set row.names
names(patClass)<-row.names(RESPdat)
# opls model
set.seed(4325)
OPLSMod<-opls(metData, patClass, subset="odd", fig.pdfC="none")
```

```
## Warning: 'permI' set to 0 because train/test partition is selected

## PLS-DA
## 105 samples x 28 variables and 1 response
## standard scaling of predictors and response(s)
##       R2X(cum) R2Y(cum) Q2(cum) RMSEE RMSEP pre ort
## Total    0.749    0.828   0.787 0.172 0.231   3   0

trainSet <- getSubsetVi(OPLSMod)

print("Fitted Model")

## [1] "Fitted Model"

table(patClass[trainSet],fitted(OPLSMod))

##
##           Resp Control
##   Resp      82       1
##   Control    0      22

print("Test Data")

## [1] "Test Data"

TestFit<-table(patClass[-trainSet],
      predict(OPLSMod, metData[-trainSet, ]))

TestFit

##
##           Resp Control
##   Resp      82       1
##   Control    3      19

TP<-TestFit[1] # True Positive
FP<-sum(TestFit[2])# False Positive
FN<-sum(TestFit[3]) # False Negative
TN<-sum(TestFit)-TP-FP-FN# True Negative

# Model of full data for plotting
pOPLSMod<-opls(metData, patClass, fig.pdfC="none")

## PLS-DA
## 210 samples x 28 variables and 1 response
## standard scaling of predictors and response(s)
##       R2X(cum) R2Y(cum) Q2(cum) RMSEE pre ort pR2Y  pQ2
## Total    0.722    0.771    0.72 0.197   3   0 0.05 0.05
```

##### Accuracy

```
(TP+TN)/(sum(TestFit))

## [1] 0.9619048
```

##### Sensitivity

```
(TP)/(TP+FN)
```

```
## [1] 0.9879518
```

##### Specificity

```
TN/(TN+FP)

## [1] 0.8636364
```

##### Plot data

NOTE: plot for training data only

```
pDat<-as.data.frame(pOPLSMod@scoreMN)
if(flipResp==T){
  pDat<-pDat*-1
}
pDat$Class<-RESPdat$OrigClass
pDat$Group<-RESPdat$Class.name
pDat$Age<-RESPdat$Age
pDat$Sex<-RESPdat$Sex
names(pDat)<-gsub("p([0-9])","Resp\\1",names(pDat))

#ggplot(aes(x=Full1,y=Full2,group=Class),data=pDat) +
#  geom_point(aes(colour=Class),size=3,alpha=0.7) + scale_colour_brewer(palette = "Set1")

#ggplot(aes(x=Full3,y=Full4,group=Class),data=pDat) +
#  geom_point(aes(colour=Class),size=3,alpha=0.7) + scale_colour_brewer(palette = "Set1")

#ggplot(aes(x=Full1,y=Full5,group=Class),data=pDat) +
#  geom_point(aes(colour=Class),size=3,alpha=0.7) + scale_colour_brewer(palette = "Set1")
```

##### Plot Control vs All Resp

```
ggplot(aes(x=Resp1,y=Resp2),data=pDat) +
  stat_ellipse(aes(colour=Group),size=1.2, alpha=0.8) +
  geom_point(aes(fill=Class,shape=Class),size=3,alpha=0.8) +
  scale_fill_manual(values=c("#989788","#E54F6D","#008BF8","#623CEA","#E7EBC5")) +
  scale_colour_manual(values=c("grey65","#E54F6D")) +
  scale_shape_manual(values=c(22,21,24,25,22))
```

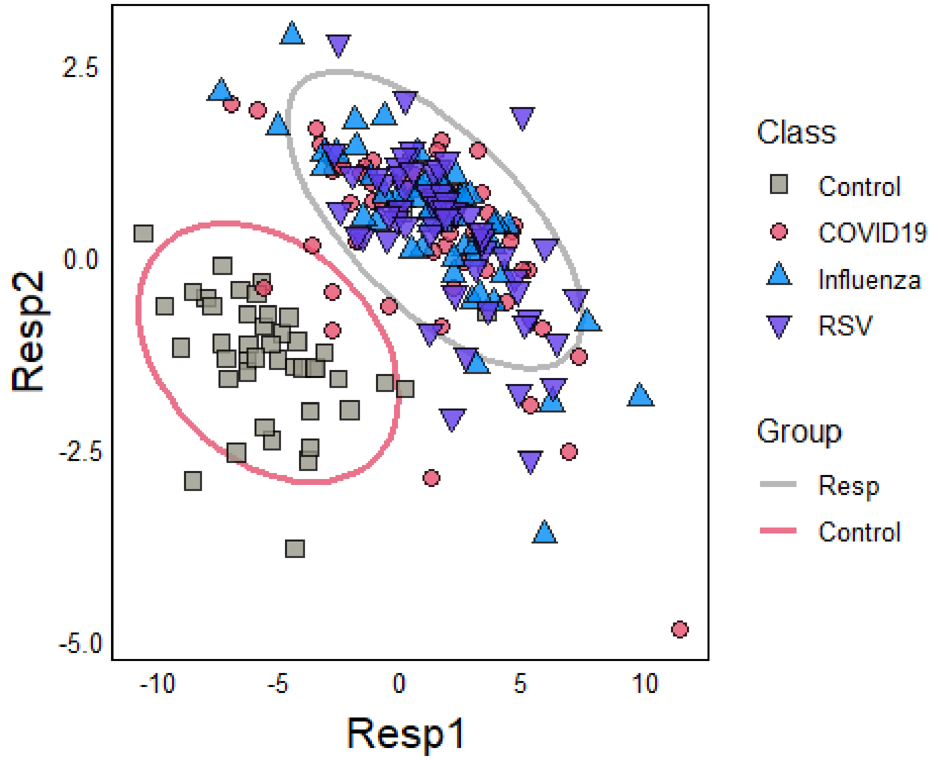


##### And the same showing age & sex

```
ggplot(aes(x=Resp1,y=Resp2,group=Class),data=pDat) +
geom_point(aes(colour=as.numeric(Age),shape=Sex),size=3,alpha=0.8) +
  scale_colour_gradient(low="blue",high="pink")

## Warning in FUN(X[[i]], ...): NAs introduced by coercion

## Warning: Removed 16 rows containing missing values (geom_point).
```

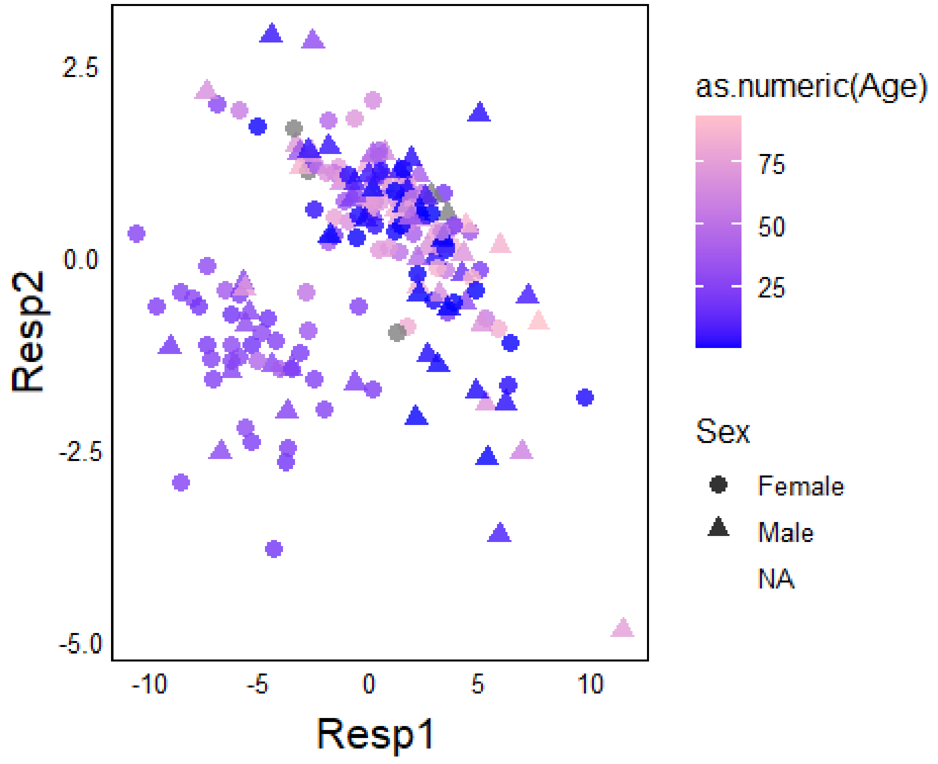

#### Export OPLS data

```
#write.csv(pDat,"./pDat/RESPdat.csv")
```

#### COVID vs Other respiratory

```
# Respiratory Only
COVIDdat<-featDatB %>% # Dataset with new encoding
    filter(Class.name %in% c("COVID19","Influenza","RSV")) %>%
    column_to_rownames("Sample.Name")
COVIDdat$OrigClass<-COVIDdat$Class.name
COVIDdat$Class.name<-gsub("Influenza|RSV","Other Resp",COVIDdat$Class.name)

DescNames<-c("Batch.Number","Class.name","Sex","Age","CT","OrigClass") # Response Variabl
e
Concs<-names(COVIDdat)[!names(COVIDdat) %in% DescNames] # Predictor Variables

# Organize data for opls
metData<-COVIDdat[,Concs] # Metabolite data
patClass<-COVIDdat[,"Class.name"] # Predictors
# Set row.names
names(patClass)<-row.names(COVIDdat)
# opls model
OPLSMod2<-opls(metData, patClass, predI = 2, subset="odd", fig.pdfC="none")

## Warning: 'permI' set to 0 because train/test partition is selected

## PLS-DA
## 84 samples x 5 variables and 1 response
## standard scaling of predictors and response(s)
##       R2X(cum) R2Y(cum) Q2(cum) RMSEE RMSEP pre ort
## Total    0.536    0.442   0.301 0.359  0.38   2   0

trainSet <- getSubsetVi(OPLSMod2)

print("Fitted Model")

## [1] "Fitted Model"

table(patClass[trainSet],fitted(OPLSMod2))

##
##              COVID19 Other Resp
##   COVID19         21          7
##   Other Resp       6         50

print("Test Data")

## [1] "Test Data"

TestFit<-table(patClass[-trainSet],
      predict(OPLSMod2, metData[-trainSet, ]))

TestFit

##
##              COVID19 Other Resp
```

```
##   COVID19         20          7
##   Other Resp       5         50

TP<-TestFit[1] # True Positive
FP<-sum(TestFit[2])# False Positive
FN<-sum(TestFit[3]) # False Negative
TN<-sum(TestFit)-TP-FP-FN# True Negative

# Model for plotting full dataset
pOPLSMod2<-opls(metData, patClass,fig.pdfC="none")

## PLS-DA
## 166 samples x 5 variables and 1 response
## standard scaling of predictors and response(s)
##       R2X(cum) R2Y(cum) Q2(cum) RMSEE pre ort pR2Y  pQ2
## Total    0.566    0.404   0.341 0.367   2   0 0.05 0.05
```

##### Accuracy

```
(TP+TN)/(sum(TestFit))

## [1] 0.8536585
```

##### Sensitivity

```
(TP)/(TP+FN)

## [1] 0.7407407
```

##### Specificity

```
TN/(TN+FP)

## [1] 0.9090909
```

##### Plot COVID vs other Respiratory

NOTE: plot for training data only

```
pDat2<-as.data.frame(pOPLSMod2@scoreMN)
if(flipCOVID==T){
  pDat2<-pDat2*-1
}
pDat2$Class<-COVIDdat$OrigClass
pDat2$Group<-COVIDdat$Class.name
pDat2$Age<-COVIDdat$Age
pDat2$Sex<-COVIDdat$Sex
names(pDat2)<-gsub("p([0-9])","COVID\\1",names(pDat2))

ggplot(aes(x=COVID1,y=COVID2),data=pDat2) +
  stat_ellipse(aes(colour=Group),size=1.2, alpha=0.8) +
  geom_point(aes(fill=Class,shape=Class),size=3,alpha=0.8) +
  scale_fill_manual(values=c("#E54F6D","#008BF8","#623CEA")) +
  scale_colour_manual(values=c("#E54F6D","grey65")) +
  scale_shape_manual(values=c(21,24,25))
```

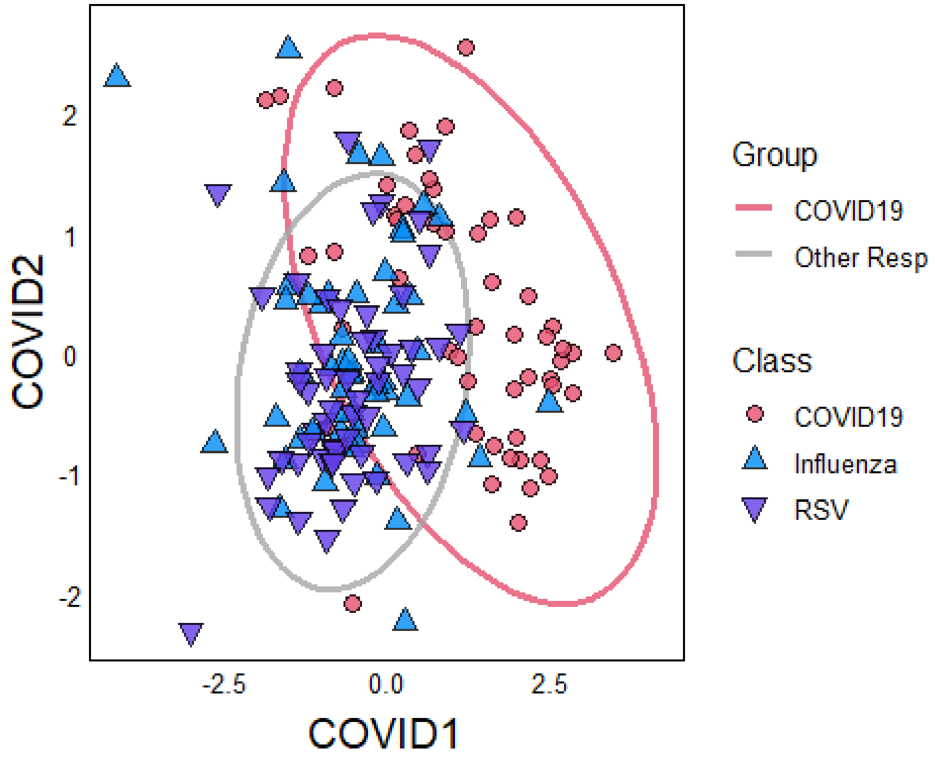


##### Age & sex

```
ggplot(aes(x=COVID1,y=COVID2,group=Class),data=pDat2) +
geom_point(aes(colour=as.numeric(Age),shape=Sex),size=3,alpha=0.8) +
  scale_colour_gradient(low="blue",high="pink")

## Warning in FUN(X[[i]], ...): NAs introduced by coercion

## Warning: Removed 16 rows containing missing values (geom_point).
```

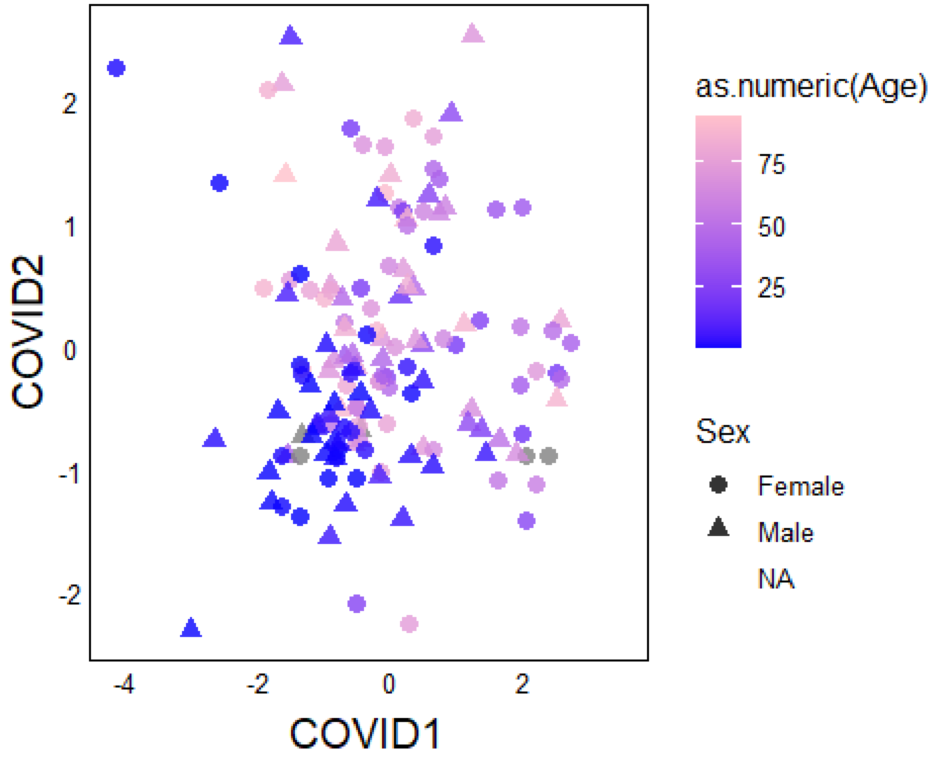

#### Export OPLS data

```
#write.csv(pDat2,"./pDat/COVIDdat.csv")
```

#### Loadings

Loading for both OPLS models

RESP = Control vs all respiratory COVID = COVID vs other respiratory

```
Loadings<-as.data.frame(OPLSMod@loadingMN)
names(Loadings)<-gsub("p","Resp",names(Loadings))
if(flipResp==T){
  Loadings<-Loadings*-1
}
cLoadings<-as.data.frame(OPLSMod2@loadingMN)
names(cLoadings)<-gsub("p","COVID",names(cLoadings))
if(flipCOVID==T){
  cLoadings<-cLoadings*-1
}
heatDat<-full_join(rownames_to_column(Loadings), rownames_to_column(cLoadings), by = "row
name")
heatDat<-gather(heatDat,Axis,Loading,all_of(names(heatDat)[-1]))
names(heatDat)[1]<-"Metabolite"
heatDat<-as.data.frame(heatDat[heatDat$Axis %in%
                                c("COVID1","Resp1"), ])

ggplot(aes(x=Axis,y=Metabolite,fill=Loading),data=heatDat) + geom_tile() +
  facet_grid(~ Axis, scales = "free_x", space = "free_x") +
  scale_fill_gradientn(colours=c("#008BF8","#E7EBC5","#E54F6D"))
```

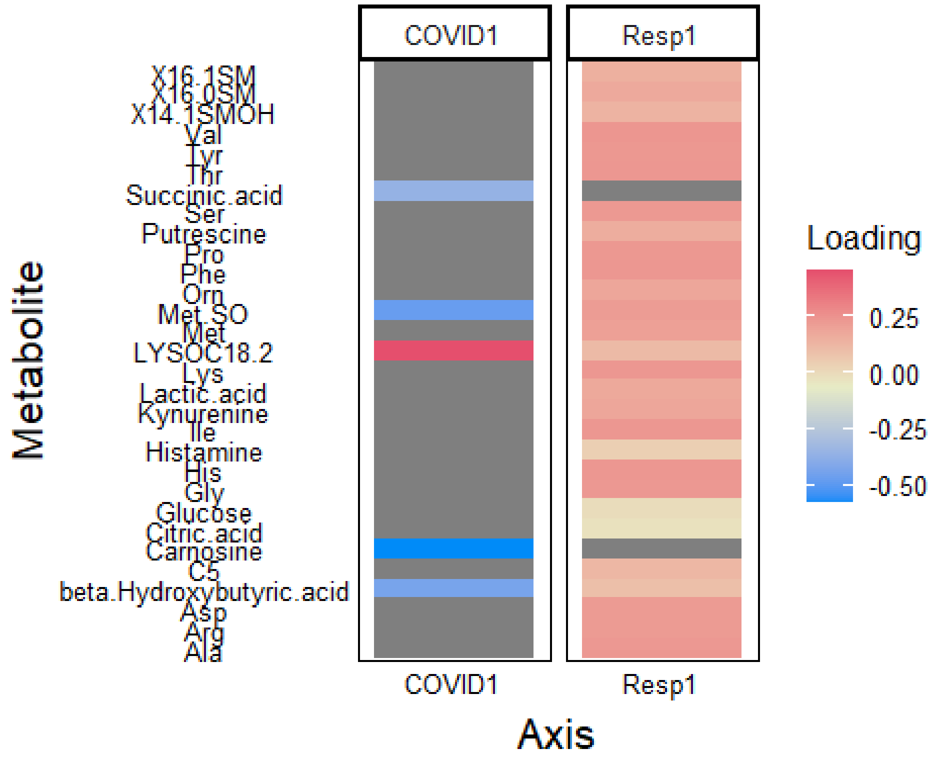

#### Export OPLS data

```
#write.csv(heatDat,"./pDat/OPLSload.csv")
```

#### Histogram of significant metabolites

```
pDat3<-gather(FULLdat,Metabolite,Concentration,
              all_of(c("Succinic.acid","Met.SO","LYSOC18.2","Carnosine","beta.Hydroxybuty
ric.acid")))
ggplot(aes(x=Concentration,group=Class.name),data=pDat3) +
  geom_density(aes(fill=Class.name),alpha=0.3) + facet_grid(Metabolite ~ .) +
  scale_fill_manual(values=c("#989788","#E54F6D","#008BF8","#623CEA","#E7EBC5"))
```

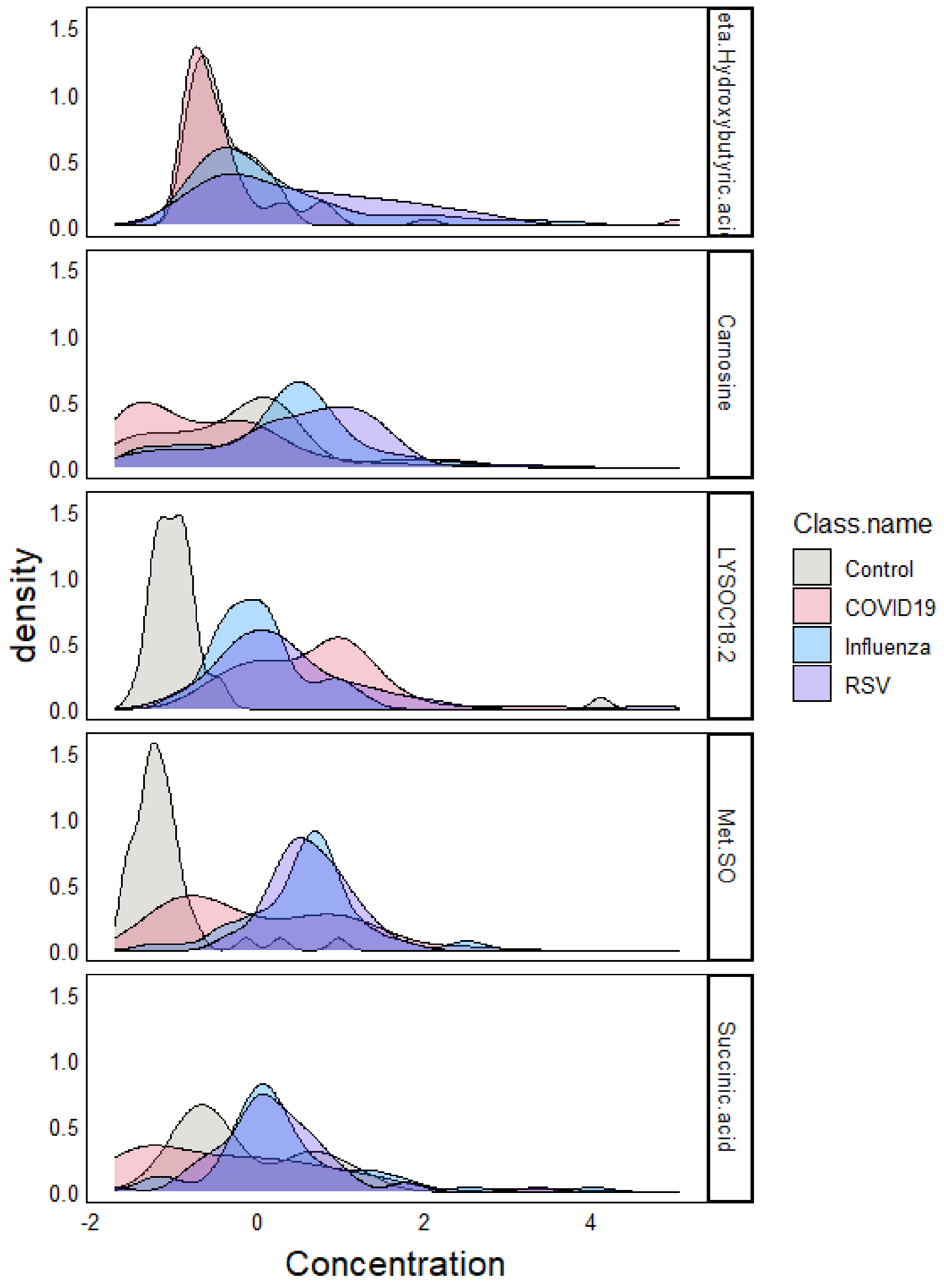


#### Export Metabolite Data

```
#write.csv(FULLdat,"./pDat/Metabolites.csv")
```

### Exploratory Data

#### CT Correlations

Do the major metabolites from COVID1 correlate with CT value in COVID and other respiratory patients?

```
COVIDmet<-c("Succinic.acid",
           "Carnosine","Met.SO","LYSOC18.2","beta.Hydroxybutyric.acid")
CTdat<-COVIDdat[,c("OrigClass","Sex","Age","CT",COVIDmet)] %>%
  gather(Metab,Conc,all_of(COVIDmet))

ggplot(aes(x=Conc,y=as.numeric(CT)),data=CTdat) +
  geom_smooth(aes(group=OrigClass,colour=OrigClass),se=F,method="lm") +
  geom_point(aes(group=OrigClass,fill=OrigClass,shape=OrigClass),
             size=3,alpha=0.8) +
  scale_fill_manual(values=c("#E54F6D","#008BF8","#623CEA")) +
  scale_colour_manual(values=c("#E54F6D","#008BF8","#623CEA")) +
  scale_shape_manual(values=c(21,24,24)) +
  facet_grid(Metab~.,scales="free")

## `geom_smooth()` using formula 'y ~ x'

## Warning: Removed 25 rows containing non-finite values (stat_smooth).

## Warning: Removed 25 rows containing missing values (geom_point).
```

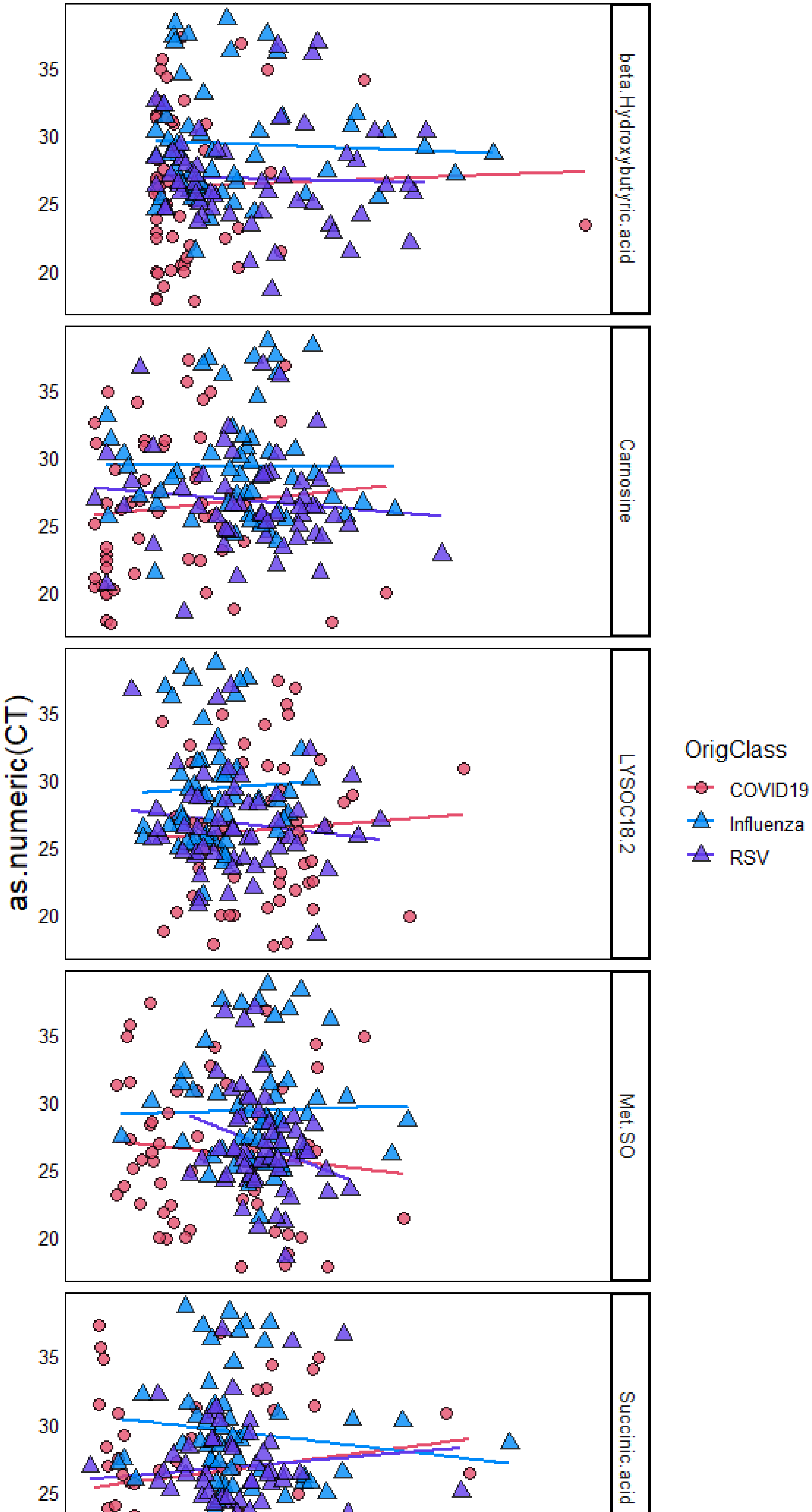
beta.Hydroxybutyric.acid
Carnosine
LYSOC18.2
Met.SO
Succinic.acid
as.numeric(CT)
20
25
30
35
OrigClass
COVID19
Influenza
RSV

#### Statistical tests

```
for(Met in unique(CTdat$Metab)){
  for(Cls in unique(CTdat$OrigClass)){
    StatDat<-CTdat[CTdat$Metab == Met &
                     CTdat$OrigClass == Cls,]
    print(anova(lm(Conc~as.numeric(CT), data=StatDat)))
    StatDat<-NA
  }
}

## Analysis of Variance Table
##
## Response: Conc
##                 Df Sum Sq Mean Sq F value Pr(>F)
## as.numeric(CT)  1  1.857   1.857  1.2914  0.261
## Residuals      52 74.775   1.438
## Analysis of Variance Table
##
## Response: Conc
##                 Df Sum Sq Mean Sq F value Pr(>F)
## as.numeric(CT)  1  0.748 0.74751  0.8974  0.348
## Residuals      50 41.649 0.83298
## Analysis of Variance Table
##
## Response: Conc
##                 Df  Sum Sq Mean Sq F value Pr(>F)
## as.numeric(CT)  1  0.2763 0.27626  0.4752 0.4936
## Residuals      53 30.8101 0.58132
## Analysis of Variance Table
##
## Response: Conc
##                 Df Sum Sq Mean Sq F value Pr(>F)
## as.numeric(CT)  1  0.325 0.32495  0.4108 0.5244
## Residuals      52 41.133 0.79101
## Analysis of Variance Table
##
## Response: Conc
##                 Df Sum Sq Mean Sq F value Pr(>F)
## as.numeric(CT)  1  0.002 0.00228  0.0029 0.9572
## Residuals      50 39.210 0.78421
## Analysis of Variance Table
##
## Response: Conc
##                 Df Sum Sq Mean Sq F value Pr(>F)
## as.numeric(CT)  1  0.612 0.61189  0.6768 0.4144
## Residuals      53 47.918 0.90411
## Analysis of Variance Table
##
## Response: Conc
##                 Df Sum Sq Mean Sq F value Pr(>F)
## as.numeric(CT)  1  0.648 0.64801  0.6642 0.4188
## Residuals      52 50.733 0.97564
## Analysis of Variance Table
##
```

```
## Response: Conc
##                 Df  Sum Sq Mean Sq F value Pr(>F)
## as.numeric(CT)  1  0.0166 0.01662  0.0333 0.8559
## Residuals      50 24.9443 0.49889
## Analysis of Variance Table
##
## Response: Conc
##                 Df Sum Sq Mean Sq F value  Pr(>F)
## as.numeric(CT)  1 0.6493 0.64928  3.5978 0.06331 .
## Residuals      53 9.5646 0.18046
## ---
## Signif. codes:  0 '***' 0.001 '**' 0.01 '*' 0.05 '.' 0.1 ' ' 1
## Analysis of Variance Table
##
## Response: Conc
##                 Df Sum Sq Mean Sq F value Pr(>F)
## as.numeric(CT)  1  0.137 0.13702  0.2065 0.6515
## Residuals      52 34.512 0.66368
## Analysis of Variance Table
##
## Response: Conc
##                 Df  Sum Sq  Mean Sq F value Pr(>F)
## as.numeric(CT)  1  0.0228 0.022824  0.0829 0.7746
## Residuals      50 13.7639 0.275278
## Analysis of Variance Table
##
## Response: Conc
##                 Df  Sum Sq Mean Sq F value Pr(>F)
## as.numeric(CT)  1  0.4731 0.47306  0.8661 0.3562
## Residuals      53 28.9470 0.54617
## Analysis of Variance Table
##
## Response: Conc
##                 Df Sum Sq Mean Sq F value Pr(>F)
## as.numeric(CT)  1  0.049 0.04939  0.0569 0.8124
## Residuals      52 45.145 0.86817
## Analysis of Variance Table
##
## Response: Conc
##                 Df Sum Sq Mean Sq F value Pr(>F)
## as.numeric(CT)  1  0.173  0.1728  0.1397 0.7102
## Residuals      50 61.866  1.2373
## Analysis of Variance Table
##
## Response: Conc
##                 Df Sum Sq Mean Sq F value Pr(>F)
## as.numeric(CT)  1  0.086 0.08638  0.0729 0.7883
## Residuals      53 62.828 1.18544
```

Stats summary: Influenze has significantly higher CT count overall, but no effect of

What about COVID1 axis from OPLS-DA model – does it predict CT values?

Setup:

```
COVIDmet<-c("Carnosine","Met.SO","beta.Hydroxybutyric.acid",
            "LYSOC18.2","Succinic.acid")
CTcomp<-COVIDdat[,COVIDmet]

CTcomp$estCOVID1<-rowSums(t(OPLSMod2@loadingMN[,1]*t(CTcomp)))

CTcomp$Sample<-rownames(CTcomp)
CTcomp$CT<-COVIDdat$CT
CTcomp$OrigClass<-COVIDdat$OrigClass
pDat2$Sample<-rownames(pDat2)

if(flipCOVID==T){
  pDat2$COVID1<-pDat2$COVID1*-1
}

pDat3<-full_join(pDat2[,c("Sample","COVID1")],
                       CTcomp[,c("Sample","estCOVID1","OrigClass","CT")],by="Sample")
```

Double-check proper calculation of COVID1 in full dataset

```
qplot(x=estCOVID1,y=COVID1,data=pDat3)
```

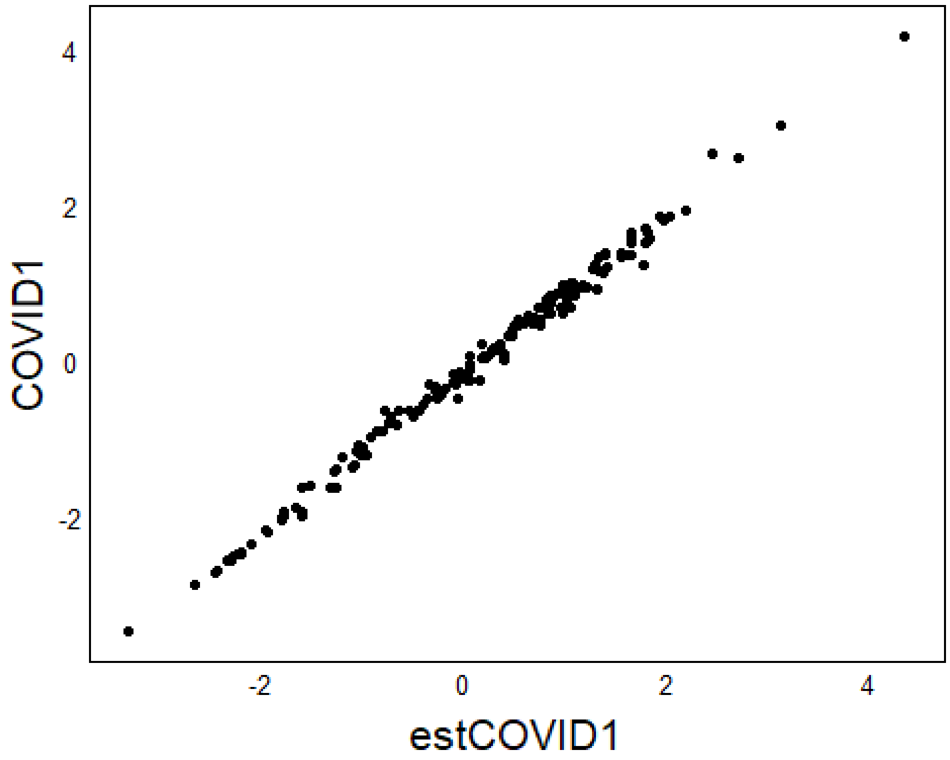


Test for CT correlation

```
ggplot(aes(x=estCOVID1,y=as.numeric(CT)),data=pDat3) +
  geom_smooth(aes(group=OrigClass,colour=OrigClass),se=F,method="lm") +
  geom_point(aes(group=OrigClass,fill=OrigClass,shape=OrigClass),
             size=3,alpha=0.8) +
  scale_fill_manual(values=c("#E54F6D","#008BF8","#623CEA")) +
  scale_colour_manual(values=c("#E54F6D","#008BF8","#623CEA")) +
  scale_shape_manual(values=c(21,24,24))
```

```
## `geom_smooth()` using formula 'y ~ x'

## Warning: Removed 5 rows containing non-finite values (stat_smooth).

## Warning: Removed 5 rows containing missing values (geom_point).
```

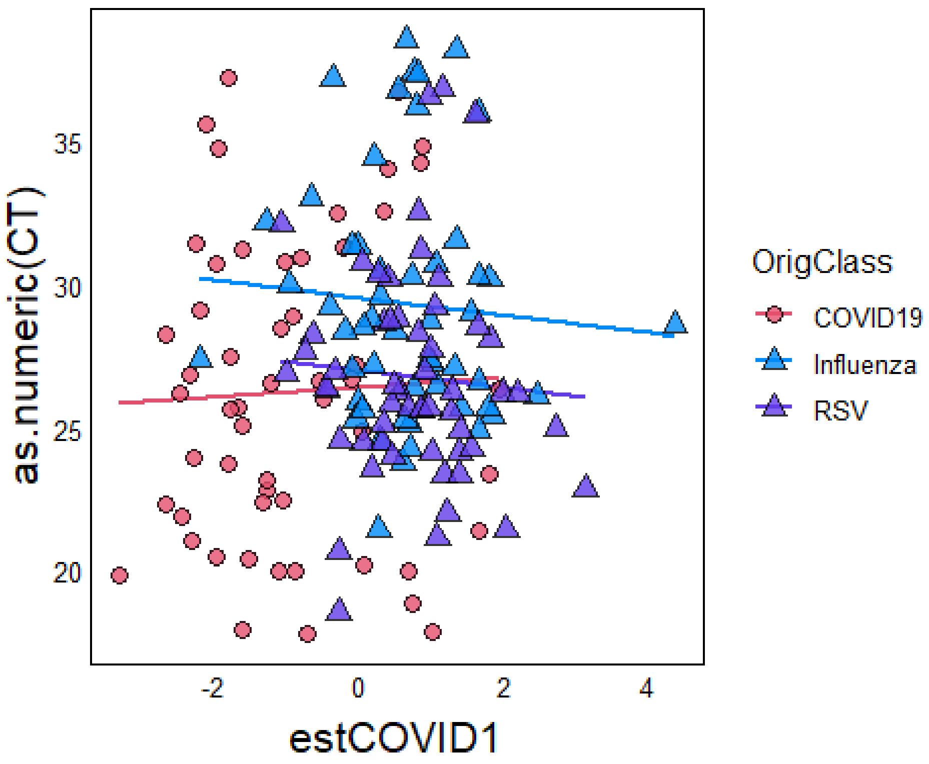
